# Persistent frustration-induced reconfigurations of brain networks predict individual differences in irritability

**DOI:** 10.1101/2021.11.07.21266032

**Authors:** J.O. Linke, S.P. Haller, E. Xu, L. Nguyen, A. Chue, C. Zapp, O. Revzina, S. Perlstein, A.J. Ross, W.-L. Tseng, P. Shaw, M.A. Brotman, D.S. Pine, S.J. Gotts, E. Leibenluft, K. Kircanski

## Abstract

**Background:** Frustration, the response to blocked goal attainment, is a universal affective experience, but how the brain embodies frustration is not known. Understanding brain network dynamics during frustration may provide insight into pediatric irritability, one of the most frequent reasons for psychiatric consultation in youth and a risk factor for affective disorders and suicidality.

**Methods:** Using fMRI, we investigated changes in neural network architecture from a baseline resting-state, through a task that included frustrative nonreward (FNR) and anticipation of new feedback following FNR (FNR+1), to a post-task resting-state in a transdiagnostic sample of 66 youth (33 female, mean age 14 years). Using a train/test/held-out procedure, we aimed to predict past-week irritability from the global efficiency (i.e., E_glob_, capacity for parallel information processing) of brain networks before, during, and after frustration.

**Results:** Compared to pre-task resting state, FNR+1 and the post-state resting state were uniquely associated with a more segregated brain network organization. Nodes that were originally affiliated with the default-mode-temporal-limbic and fronto-parietal networks contributed most to this reconfiguration. Solely E_glob_ of brain networks that emerged after the frustrating task predicted self- and observer-rated irritability in previously unseen data. Self-reported irritability was predicted by E_glob_ of a fronto-temporal-limbic module, while observer-rated irritability was predicted by E_glob_ of motor-parietal and ventral-prefrontal-subcortical modules.

**Discussion:** We characterize frustration as an evolving brain network process and demonstrate the importance of the post-frustration recovery period for the pathophysiology of irritability; an insight that, if replicated, suggests specific intervention targets for irritability.

## Introduction

Irritability is a common reason for pediatric psychiatric consultation and a risk factor for adult psychopathology (1) and suicidality (2). Aberrant responses to frustration are thought to be a key mechanism of irritability (3). Frustration is a complex emotional and motivational state associated with distributed brain regions, but brain network dynamics related to the emergence and recovery from frustration are poorly understood; such knowledge could guide the development of targeted interventions for irritability. Here, we coupled fMRI during a frustrating task with pre- and post-task resting state scanning to characterize frustration as an evolving process from a brain network perspective. Further, we probe the utility of network metrics for predicting irritability.

Frustration occurs when actions fail to yield an expected reward (i.e., frustrative non-reward; FNR; 4). Three studies have investigated brain activation during or after FNR in healthy adults. These found effects in widely distributed cortical and subcortical brain regions (5–7), highlighting the need for circuit-based approaches. FNR also evokes anger and aggression (8). In twenty-one traumatized males, anger induction was associated with increased amygdala-inferior frontal gyrus connectivity during a subsequent resting-state scan, (9) emphasizing the need for in-depth analysis of the chronometry of FNR-induced brain network changes.

Studies using frustration tasks in youth have focused on clinical samples enriched for irritability. These studies associate irritability with aberrant activity of widely distributed brain regions during FNR (10,11), attention orienting post-FNR (12), and reduced youth-caregiver prefrontal synchrony post-FNR (13). Recently, individual differences in irritability were predicted from large-scale brain network connectivity during frustrating task blocks (14). Together, these findings suggest the relevance of functional brain networks before, during, and after frustration in elucidating the mechanisms of irritability.

Reconfiguration of functional brain networks in response to different contexts (rest, frustration) and stimuli can be studied using a graph-theoretical approach. This framework assumes that the brain is organized into complex subnetworks (modules) consisting of nodes (brain regions) and edges (functional connectivity between regions; 15,16). Investigators can calculate a global measure of the brain’s organization into non-overlapping subnetworks (modularity) to characterize the nodal composition of these modules and examine the capacity for parallel information processing within modules (efficiency; 16).

Graph-theoretical approaches have been used to understand the brain at rest and, to a lesser extent, during cognitive tasks (17). Few studies have used this approach to examine changes in brain networks in response to affective stimuli. One found that cues signaling possible reward or shock elicited shifts toward a more integrated modular organization (18), while another found that trait emotional expression was related to fronto-parietal network (FPN) and default mode network (DMN) efficiency (19).

Here, we investigated frustration from a brain network perspective, comparing global (modularity/brain network segregation, Aim 1a) and local (modular composition, Aim 1b) brain network properties before, during and after a frustrating task. Further, we tested the predictive value of brain module characteristics (global efficiency) for self-rated frustration, and trait irritability (Aim 2).

## Methods and Materials

### Sample

We recruited a transdiagnostic sample that was racially, ethnically and socioeconomically diverse and enriched for irritability (i.e., youth with disruptive mood dysregulation disorder, attention-deficit/hyperactivity disorder and/or an anxiety disorder and healthy youth; N=66, 33 female, mean age=14.0 years). Diagnoses were made using the Kiddie Schedule for Affective Disorders and Schizophrenia (20). We controlled for effects of psychotropic medication (n=25) by covarying for medication load (21). Exclusion criteria were neurological, autism spectrum, and bipolar disorders, psychosis, substance use, MRI contraindications, and IQ<70. Consent, assent and Institutional Review Board approval were obtained.

### Paradigm

Participants underwent a 9-minute resting-state scan, then a 14-minute frustration-inducing, attention-orienting task (100 trials over two runs), followed by another 9-minute resting-state scan. During the modified Posner task, participants press a button as fast as possible to indicate a target location after a valid (75%) or invalid (25%) cue. To establish reward expectation before scanning, participants completed a task version (Game 1) in which correct responses (∼98% of trials) were rewarded with $.50. During scanning, frustation was added: after 60% of correct responses, $.50 were deducted under the pretense that responses were too slow (Game 2; **Figure 1**; 10,12).

**Figure 1.**
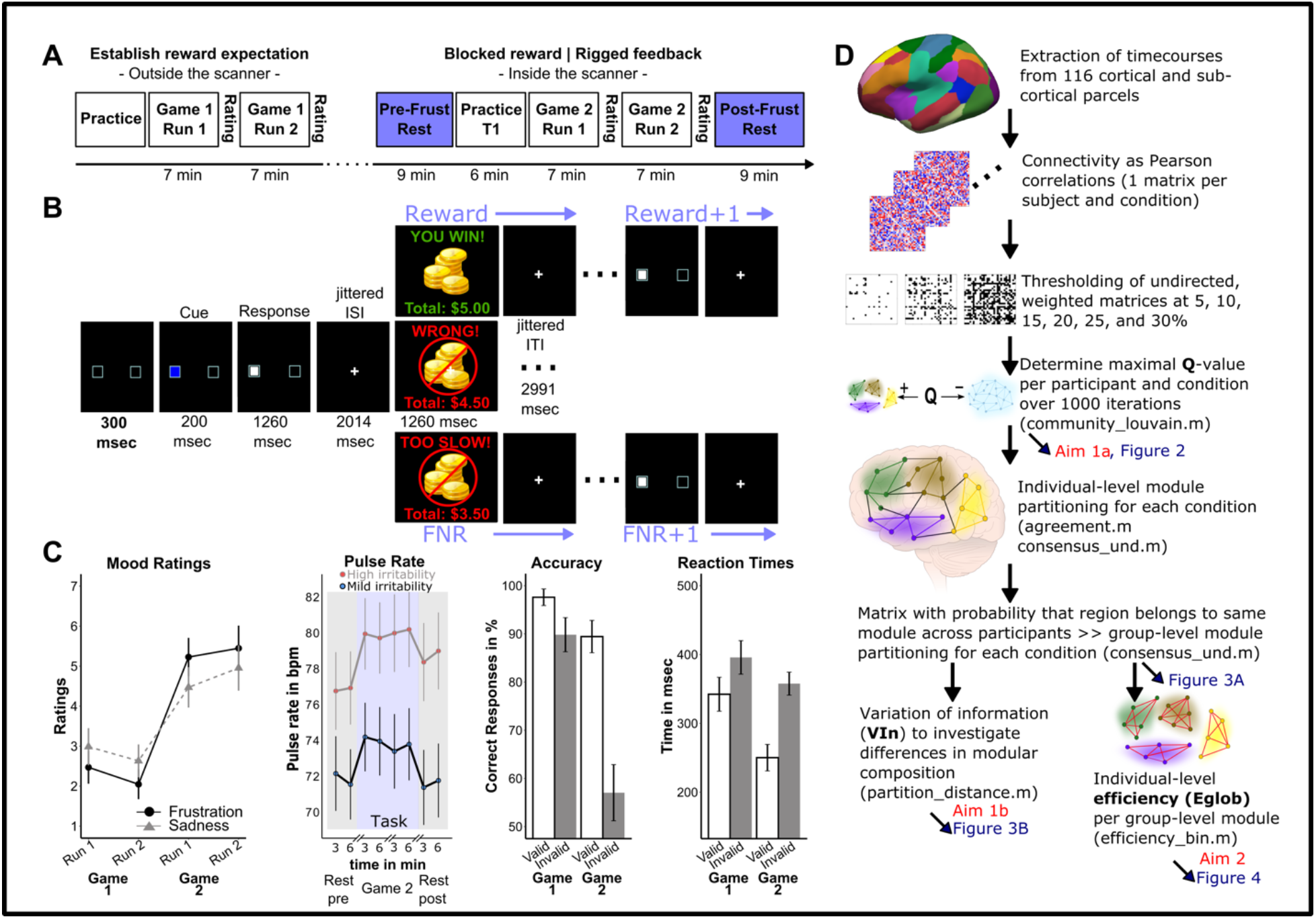
Experimental design, trial sequence, behavioral results, and processing pipeline. **Panel A** provides an overview of the paradigm. Game 1, conducted outside the scanner, established a reward expectation. Game 2 induced frustration during functional magnetic resonance imaging. Before and after Game 2, nine minutes of resting-state data were acquired. **Panel B** shows order and timing of one trial. Feedback can be FNR (frustrative non-reward) or Reward. FNR+1 denotes the anticipation phase on trial N+1, where the feedback on trial N was FNR. Similarly, Reward +1 denotes the anticipation phase on trial N+1, where the feedback on trial N was Reward. **Panel C** shows several behavioral measures. We see the course of mood ratings (frustration and sadness) before (Game1) and during (Game 2) frustration. Pulse rate was recorded during fMRI only. Thus, here we see it’s course during first and last half of pre-task resting state, run 1, run 2, and post-task resting state as a function of parent-rated irritability. It also shows the Posner-effect (lower accuracy and longer reaction times to invalid vs. valid cues) across Games 1 and 2. Error-bars represent 95% confidence intervals. **Panel D** provides an overview of the processing pipeline. It shows the main processing steps, how the three metrics of interest (Q, VIn, Eglob) were acquired, and how they relate to study aims and figures. The scripts from the Brain Connectivity Toolbox that were used during the different steps are also noted. Condition is used here as an umbrella term that referes to the pre- and post-task resting-state and the four events of interest. **Abbreviations: *bpm***, beats per minute; ***Eglob***, global efficiency; ***FNR***, frustrative non-reward; ***Frust***, frustration; ***ITI***, inter-trial interval; ***ISI***, inter-stimulus interval; ***msec***, milliseconds, ***Q***, modularity index Q; ***VIn***, Variation of information

We modeled brain network configurations during six conditions; pre- and post-task resting-state (pre-RS and post-RS, respectively) and four task events: (1) frustrating deduction of $0.50 (FNR), (2) feedback anticipation during trials immediately following FNR (FNR+1), (3) winning $0.50 (Reward), and (4) feedback anticipation during trials immediately following Reward (Reward+1). Reaction time (RT) and accuracy were measured for valid trials, where the cue correctly predicts the target location, and for cognitively more demanding invalid trials. Participants rated their frustration after each run. Pulse rate (PR) from finger photoplethysmography (22) was recorded throughout scanning. Increased PR reflects physiological arousal (23) associated with (24), albeit not specific to, frustration (25). Before scanning, parents and youth independently rated the child’s irritability during the past week with the Affective Reactivity Index (26). Behavioral data were analyzed using repeated-measures analysis of variance (ANOVA); linear mixed effects models were used for PR (for details, see **Supplement**).

### Preprocessing of imaging data

Functional and structural imaging data were acquired on two identical 3.0 Tesla scanners. Data quality was assessed using MRIQC (v0.15.2; 27). Fourteen participants were excluded for motion-related artifacts (framewise displacement >0.5mm/repetition time for > 30% of the images). Data from the remaining 66 participants were preprocessed with FMRIPREP (v20.0.5; 28).

### Network construction

The functional connectivity network comprised 116 nodes: 100 cortical parcels assigned to known functional networks (29) and 16 subcortical regions from the FMRIPREP FreeSurfer segmentation. We regressed out from the time series motion parameters, ICA-AROMA head motion components, white matter and CSF signal, the first three principal components from aCompCor, the first three cosine variables, framewise displacement and the spatial standard deviation of the temporal difference data.

Task conditions convolved with a canonical hemodynamic response function were regressed from the time series to remove variance associated with task-related coactivation (30,31). After accounting for the hemodynamic lag, we created a time series specific to each task event for each node by concatenating the residual time series associated with the relevant events within and across the two runs. Functional connectivity was quantified using Pearson correlations transformed for normality using Fisher’s *z’* transformation (**Figure 1D**).

### Aim 1a: Does brain network segregation differ among the six conditions?

First, we calculated the modularity index (Q) for each condition (32). Higher Q-values indicate a network partition with many connections within and few between modules, defined as non-overlaping groups of nodes (33), and thus more localized, segregated information processing (**Figure S2**). Q was estimated using the Louvain greedy algorithm (34) implemented in the Brain Connectivity Toolbox (16). Given the stochastic initialization of the greedy optimization, it was applied 1000 times for each condition. The highest Q-value was used to compare modularity indices among conditions with paired t-tests using 5000 permutations, threshold of p<.05 applying family-wise error rate (FWER) correction across density thresholds (5%, 10%,15%, 20%, 25%, 30%), (35) and Hedges’ *g* effect size. We ensured consistent results across different graph density thresholds, but focus on the description of results obtained using a 10% threshold. Modularity shifts during task were entered as covariates into the repeated-measures ANOVAs of the behavioral data.

### Aim 1b: Does the modular composition of brain networks differ among pre-task resting state, task events and post-task resting state?

Second, we characterized the brain networks specific to the six conditions. Multiple module partitions maximize Q. Thus, we used a consensus approach to calculate an agreement matrix across the 1000 iterations for each participant and condition. For each condition, we calculated a matrix reflecting the probability of nodes being assigned to the same module across participants (31) and subjected these matrices to the same community detection algorithm used at the individual level. Across conditions, we used the variation of information metric (VIn; 36) to quantify the degree of dissimilarity in their modular composition. Significant differences in modular structure were determined using a repeated-measures permutation procedure with 5000 permutations (31,37). To determine the contribution of specific modules to significant overall reconfiguration, we compared the VIn-values for each module between conditions using paired t-tests with 5000 permutations (31).

### Aim 2: Does information processing efficiency of specific modules during any of the six conditions predict task performance, frustration, and/or irritability?

Lastly, we tested whether the global efficiency (E_*glob*_) of any module during the six conditions predicted task performance, task-induced changes in frustration, or irritability. Prior work associates E_*glob*_ positively with neurophysiological (38,39), cognitive (31,40), and emotional functioning (19). It is defined as the inverse of the average path length between all nodes (41) and indexes the capacity for parallel information processing within a module. We calculated E_*glob*_ within each module from the group-level modularity partition at 10% network density. We used a prediction framework, dividing the sample into training/validation and held-out/testing subsets (80/20) using stratified random sampling. Predictors comprised the efficiency of all modules during the six conditions, mean framewise displacement specific to each condition, age, sex, medication load, and scanner. In the training/validation dataset, predictors were selected using linear stepwise regression, applying a 10-fold cross-validation with 20 repeats as implemented in the Caret package for R. The resulting model was used to predict task performance, change in frustration ratings, youth- and parent-rated irritability in the held-out dataset. We used 5000 permutations and a threshold of p<.05 applying False Discovery Rate (FDR) correction across the five models. To determine specificity, we tested whether task-induced increase in sadness and symptom-ratings of anxiety, inattention, and hyperactivity could be predicted. A more detailed description of the methods can be found in the **Supplement**.

## Results

### Task performance, frustration ratings, pulse rate, and irritability

As expected, invalid cues were associated with longer RT (F_(1,63)_=20.66, p<.001, η^2^=.25) and lower accuracy (F_(1,63)_=22.41, p<.001, η^2^=.27). The introduction of FNR (Game 2), was associated with faster responses during valid trials (F_(1,63)_=9.60, p=.003, η^2^=.13), more errors during invalid trials (F_(1,63)_=7.95, p=.006, η^2^=.11), and increased frustration compared to Game 1 (F_(1,63)_=18.26, p<.001, η^2^=.23, **Figure 1C**). PR was best modeled as a quadratic function of time (i.e., peaking during the task) with subject as a random effect. Because age (β=−.99, t_(59.2)_=−2.15, p=.036) and parent-rated irritability (β=3.66, t_(56)_= 2.83, p=.006) predicted PR, they were included as fixed effects (**Supplement)**. Steeper PR increase during the task was associated with more errors on invalid trials (F_(2,52)_=3.33, p=.001, η^2^=.11).

### Aim 1a: Does brain network segregation differ among the six conditions?

Compared to pre-RS, the brain transitioned into a more global processing mode during task events (Reward: t_(65)_=−6.51, p_FWER_<.001, Hedges’g=−1.04; Reward+1: t_(65)_=−7.07, p_FWER_<.001, Hedges’g=−1.31; FNR: t_(65)_=−4.10, p_FWER_<.001, Hedges’ g=−0.60). There was one exception: FNR+1 elicited more localized processing (t_(65)_=6.93, p_FWER_<.001, Hedges’ g=1.36). Post-RS was also associated with more localized processing (t_(65)_=2.88, p_FWER_=.008, Hedges’s g=0.48), although the difference from pre-to post-RS was less pronounced than from pre-RS to FNR+1 (**Figure 2A, S3**). There were no associations between modularity and motion (**Table S2**).

**Figure 2.**
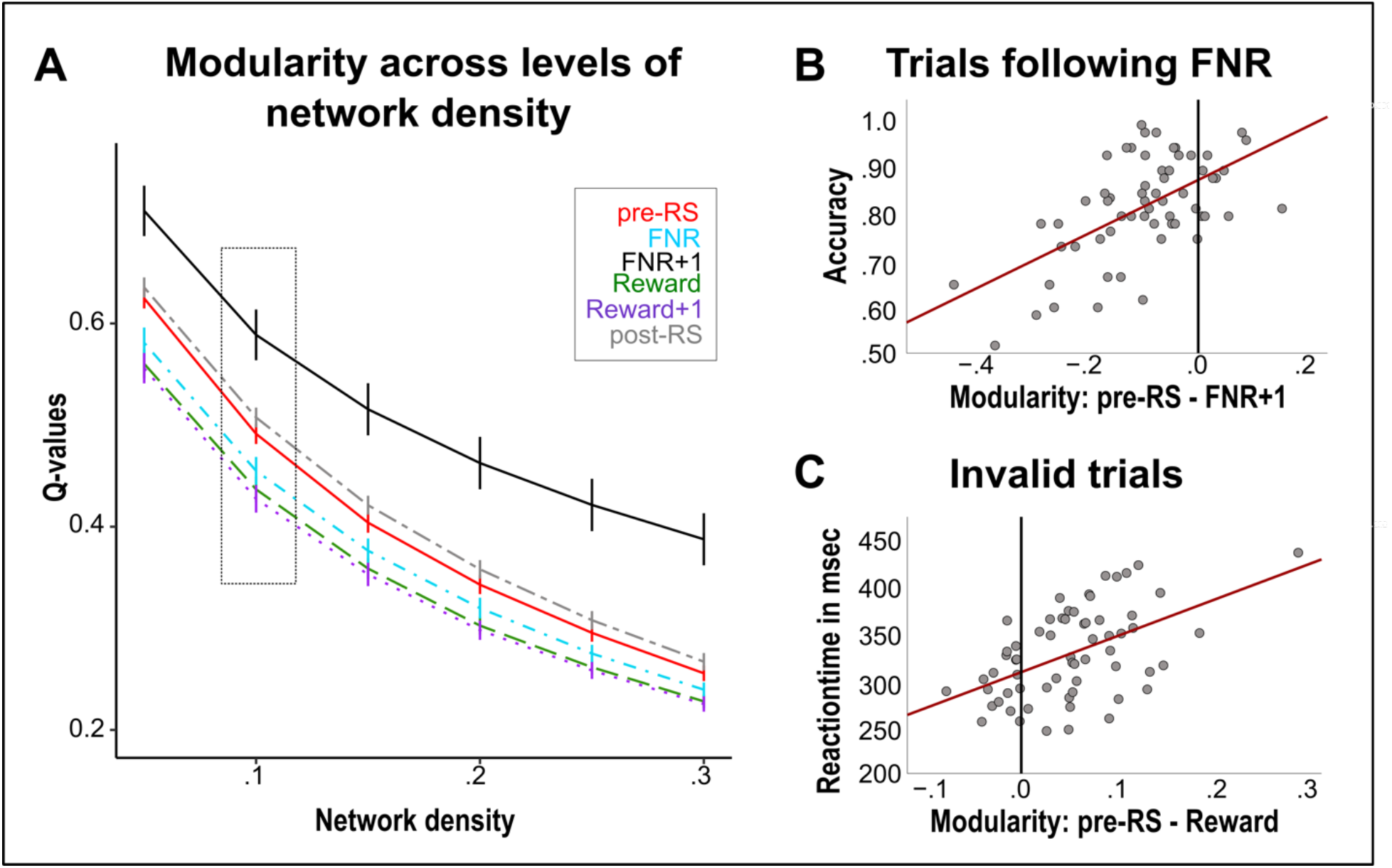
Changes in modularity (Q) throughout the paradigm and associations with behavior. **Panel A** shows that during Reward, Reward+1, and FNR the brain transitioned to a more global processing mode relative to the pre-task resting state. However, during FNR+1 and the post-task resting state period, functional segregation of the brain was higher. This pattern is present across network densities shown on the x-axis. **Panel B** illustrates how the shift towards a more localized processing mode during FNR+1 relates to lower accuracy in trials following FNR. **Panel C** depicts how the transition towards a more local processing mode during Reward is associated with faster reaction times in invalid trials. **Abbreviations: *post-RS***, post-task resting state; ***pre-RS***, pre-task resting state

Larger shifts towards a more global processing mode during Reward (F_(53,1)_=6.78, p=.012, η^2^=.12, **Figure 2C**) and at trend-level during FNR (F_(53,1)_=3.83, p=.057, η^2^=.08) were associated with longer RT in the cognitively more demanding invalid trials. Larger shifts towards a more localized processing mode during FNR+1 were associated with lower accuracy during Game 2 (Game×Shift: F_(1,54)_=5.23, p=.026, η^2^=.09, **Figure** 2B), and less RT slowing on invalid trials following FNR (Validity×Feedback×Shift: F_(1,53)_=11.80, p=.001, η^2^=.19).

### Aim 1b: Does the modular composition of brain networks differ among the six conditions?

Pre-RS included seven modules (**Figure 3A**). Based on the Schaefer atlas and previous literature (30), we labeled these modules visual (VIS), cingulo-opercular (CO), parietal (PAR), fronto-parietal (FP), anterior DMN-temporal-limbic (aDMN-TL), salience (SAL) and subcortical (SC; **Figures S4-S10**).

**Figure 3.**
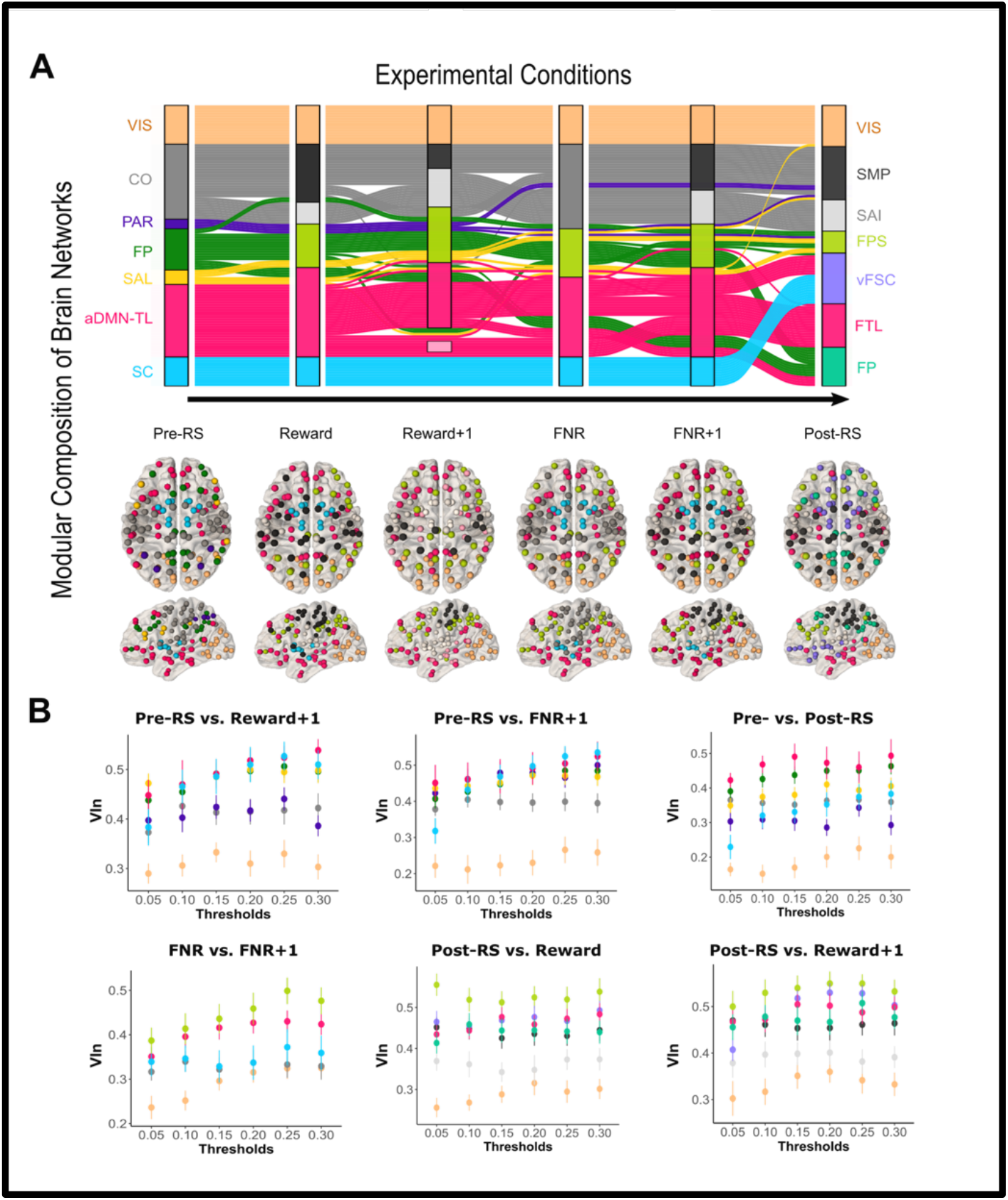
Modular reconfiguration of the whole-brain network during the six experimental conditions. The alluvial diagram in **Panel A** illustrates the reconfiguration of brain network modules from the pre-task resting state (left) through the four task conditions (i.e., Reward, Reward+1, FNR, FNR+1) and the post-task resting state (right). Modules identified in each condition are shown in the vertical boxes. During Reward+1 several nodes of the aDMN-TL and SC modules could not be assigned to modules with more than three nodes. The height of the boxes corresponds to the number of nodes within each module and the streamlines depict how nodes originally belonging to one network change their membership throughout the paradigm. Below the flow diagram, nodes are overlaid on a brain template with colors corresponding to the module to which they belonged during that condition. **Panel B** shows the VIn values across the different modules for the task conditions, where significant differences between conditions were observed. The color scheme of the nodes is consistent with the one used in the alluvial plot for the first condition mentioned in the title of each plot. Error bars represent 95% confidence intervals. **Abbreviations: *aDMN-TL***, anterior default mode network plus temporal and limbic networks**; *CO***, cingulo-opercular module; ***FNR***, frustrative non-reward (i.e., rigged feedback); ***FNR+1***, anticipation of new feedback after rigged feedback in previous trial; ***FP***, fronto-parietal module; ***FPS***, fronto-parietal-salience module; ***FTL***, fronto-temporal-limbic module; ***PAR***, parietal module; ***post-RS***, post-task resting state; ***pre-RS***, pre-task resting state; ***Reward+1***; anticipation of new feedback after rewarding feedback in previous trial; ***SAI***, somato-auditory-insular module; ***SAL***, ventral salience attention module; ***SC***, subcortical module; ***SMP***, somato-motor-parietal module; ***VIn***, variation of information; v***FSC***, ventral fronto-subcortical module; ***VIS***, visual module

Across network densities, the modular composition during pre-RS changed in response to Reward+1 (VIn=.30, p<.001), FNR+1 (VIn=.18, p<.001), and post-RS (VIn=.22, p<.001; **Table S1**). We also observed differences between FNR and FNR+1 (VIn=.10, p=.002). Modular composition during FNR did not differ from pre-RS (VIn=.12, p=.78). The effect of Reward (VIn=.16, p=.009) was unstable (**Table S1**).

We determined the relative contribution of each module to differences in overall modular composition by calculating the VIn for individual modules (31). A relatively higher VIn value indicates a greater contribution of that module to the overall reconfiguration. VIS showed the lowest VIn-values (all p_FWE_<.0001, **Figure 3B**), while aDMN-TL and FP contributed most to the reconfigurations. Comparing pre-RS to both FNR+1 and Reward+1, aDMN-TL (all p_FWE_<.0003) and FP (all p_FWE_<.0072) showed higher VIn than CO. Differences between FNR and FNR+1 were also driven by aDMN-TL and FP, which showed higher VIn-values than all other modules (all p_FWE_<.0483). Differences between pre- and post-RS were also driven by aDMN-TL and FP, which showed higher VIn than CO (all p_FWE_<.0019), PAR (all p_FWE_<.0399), and SC (all p_FWE_<.0300, **Figure 3B**).

Visual inspection indicated that during the task, one part of the FP branched off to merge with the aDMN-TL, while the remaining nodes were joined by nodes originally affiliated with PAR and SAL. Further, CO split into two parts, one joining the somato-motor-parietal (SMP) and one joining the somato-auditory-insula (SAI) part during Reward, Reward+1 and FNR+1 (**Figure 3A**). During post-RS, two conjoined modules emerged. Specifically, the ventral prefrontal nodes of aDMN-TL merged with SC to form the ventro-frontal-subcortical module (vFSC); the remaining nodes will be referred to as fronto-temporal-limbic module (FTL). In addition, nodes originally affiliated with the FP, PAR and SAL merged to form a fronto-parietal-salience module (FPS). (**Figures 3A, S5-S10)**. SMP and SAI were also found during post-RS.

Does frustration drive differences between pre- and post-task network reconfiguration? We reasoned that modular composition differences between post-RS and both Reward and Reward+1, coupled with the absence of differences between post-RS and both FNR and FNR+1, would provide evidence, albeit not proof, that this is the case. Indeed, we observed differences among post-RS, Reward (VIn=.26, p=.005) and Reward+1 (VIn=.36, p<.001) that were driven by the post-RS FPS (all p_FWE_<.0493, **Figure 3B)**. No differences in modular composition between post-RS and either FNR (VIn=.26, p=.525) or FNR+1 (VIn=.23, p=.960) were observed, supporting the hypothesis that frustration drives differences between pre- and post-RS.

### Aim 2: Does information processing efficiency of specific modules during any of the six conditions predict task performance, frustration, and/or irritability?

In the training/validation dataset, age (β=0.41) and FP E_*glob*_ during pre-RS (β=−0.29) and FNR+1 (β=0.41) predicted the RT difference between valid and invalid trials. Age (β=0.34), aDMN-TL (β=-0.29), and SC (β=0.26) E_*glob*_ during FNR+1 predicted accuracy. These models also predicted task performance in the held-out dataset, although marginally for accuracy (RT: r=.59, p_FDR_=.011, R^2^=.35, RMSE=0.93; accuracy: r=.46, p_FDR_=.054, R^2^=.21, RMSE=1.01).

In the training/validation dataset, participants’ increase in frustration was predicted by baseline frustration ratings (β=-0.29), VIS (β=−0.28) and aDMN-TL E_*glob*_ during Reward (β=−0.23) and FTL E_*glob*_ during post-RS (β=0.25, **Figure 4**); these were also predictive in the held-out subset (r=.62, p_FDR_=.005, R^2^=.38, RMSE=0.81).

**Figure 4.**
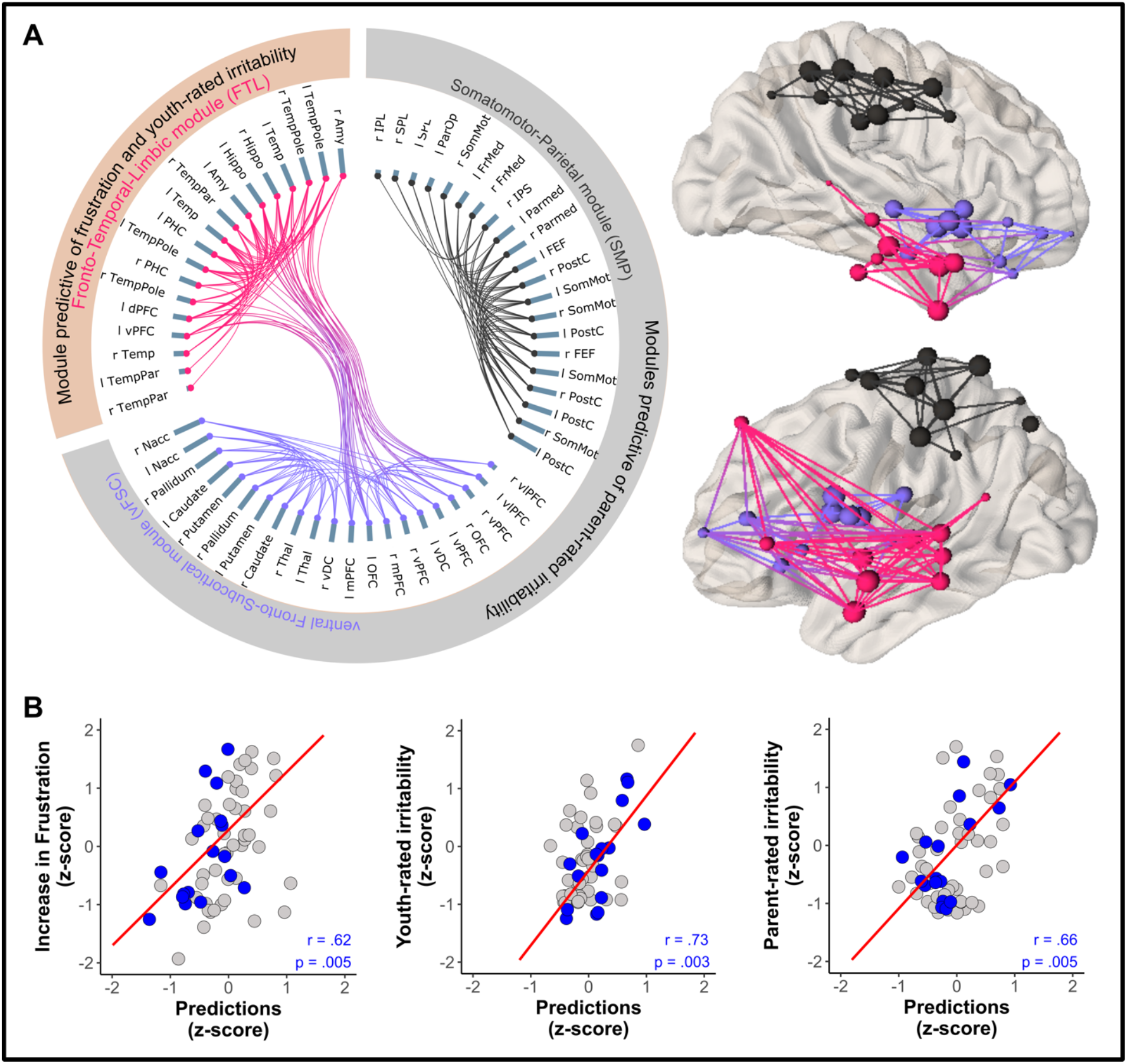
Module efficiency as predictor of frustration, youth- and parent-rated irritability. **Panel A** shows the modules that emerged during the post-task resting state and predicted either increase in frustration and youth-rated irritability (pink FTL module) or parent-rated irritability (grey SMP and lilac vFSC modules). The length of the grey-blue bars for each node represents the centrality of this node within the module, a graph-theoretical measure of the closeness of this node relative to all the other nodes in this module. In separate graphs, **Panel B** illustrates associations between the predicted increase in frustration, youth-rated irritability and parent-rated irritability vs. the actual values in the held-out dataset (blue dots). Pearson correlation coefficients and corrected p-values are also shown. This is overlaid on the predictions of the regression model for the training/validation dataset (grey dots). **Abbreviations: *Amy***, Amygdala; ***dPFC***, dorsal prefrontal cortex; ***FEF***, frontal eye field; ***FrMed***, medial frontal cortex; ***Hippo***, Hippocampus; ***IPL***, inferior parietal lobe; ***IPS***, intraparietal sulcus; ***L***, left; ***mPFC***, medial prefrontal cortex; ***Nacc***, nucleus accumbens; ***OFC***, orbitofrontal cortex; ***ParMed***, medial parietal cortex; ***ParOper***, parietal operculum; ***PHC***, parahippocampal cortex; ***R***, right; ***SomMot***, somato-motor cortex; ***SPL***, superior parietal lobe; ***Temp***, temporal; ***TempPar***, temporoparietal region; ***TempPole***, temporal pole; ***ventral DC***, ventral diencephalon, which includes hypothalamus, mammillary bodies, subthalamic nuclei, substantia nigra, red nucleus, and medial and lateral geniculate nuclei; ***vPFC***, ventro-medial prefrontal cortex; ***vlPFC***, ventro-lateral prefrontal cortex

In the training/validation dataset, post-RS FTL E_*glob*_ (β=-0.25) and medication load (β=0.38) predicted youth-rated irritability; these were also predictive in the held-out subsample (r=.73, p_FDR_=.003, R^2^=.53, RMSE=0.73, **Figure 4**). Post-RS SMP (β=0.38) and vFSC (β=0.40, **Figure 4**) E_*glob*_ predicted parent-rated irritability; these were also predictive in the held-out dataset (r=.66, p_FDR_=.005, R^2^=.44, RMSE=0.82). Models predicting task-induced increases in sadness, changes in PR, anxiety, inattention, and hyperactivity in the training/validation subset showed no utility in previously unseen data (all p_uncorrected_>.13).

## Discussion

We report three key findings. First, compared to pre-RS, the two psychological conditions that represent adaptations to, and recovery from FNR (i.e., FNR+1, post-RS) were associated with (a) more localized processing, and (b) changes in modular composition driven largely by nodes originally affiliated with the FP and aDMN-TL modules. Second, similar modular composition between FNR+1 and post-RS, coupled with differences between post-RS and non-frustrating task conditions, suggest that frustration drives differences between pre- and post-RS. Finally, E_*glob*_ of post-RS modules predicted task-induced increases in frustration-ratings and irritability, underscoring the relevance of the recovery from frustration for the pathophysiology of irritability.

### Aim 1a: Does brain network segregation differ among the six conditions?

Consistent with prior work associating threat- and reward-cues with lower brain network modularity (18), we observed reduced brain network segregation during Reward, Reward+1 and, to a lesser degree, FNR relative to pre-RS. More global processing during Reward and FNR was associated with longer RTs during the cognitively demanding invalid cue condition. In contrast, during FNR+1, we observed increased brain network segregation related to shorter RTs, and lower accuracy i.e., a speed-accuracy trade-off (42). A more localized processing mode also characterized post-RS, relative to pre-RS. Future studies should examine the duration of network reconfiguration following frustration, and potential effects on psychological processes.

### Aim 1b: Does the modular composition of brain networks differ among the six conditions

Across conditions, reconfiguration of brain networks were driven by nodes originally affiliated with FP and aDMN-TL. These modules comprise regions previously implicated in frustration (FP: posterior cingulate, precuneus; aDMN-TL: ventral and middle PFC, amygdala; 5–7) Multiple studies suggest that FP (31,37,43) and aDMN-TL (44,45) nodes play a crucial role in adaptating to environmental demands. This view is supported by our data associating FP and aDMN-TL configuration with performance during attention reorientation.

Our findings also highlight the role of frustration in brain network reconfigurations. Post-RS brain configuration differs from the two reward-related events, but not the two FNR-related events. This suggests that the frustrating aspects of the task drive differences between pre- and post-RS. Moreover, pre-RS differed from FNR+1 and post-RS, two conditions relevant for the recovery from FNR. Together, these findings underscore the relevance of recovery from FNR for understanding frustration and mental disorders characterized by prolonged angry mood (9,13).

### Aim 2 Does information processing efficiency of specific modules during any of the six conditions predict task performance, frustration, and/or irritability?

Using a train/test/held-out procedure, we predicted task-induced increases in frustration and parent- and youth-rated irritability. During post-RS, nodes originally affiliated with aDMN-TL were key for these predictions. Specifically, during post-RS, ventral prefrontal nodes of aDMN-TL merged with SC nodes and the resulting vFSC module predicted parent-rated irritability, while the remaining FTL module, consisting of frontal, temporal and limbic regions, predicted youth-rated measures of both frustration and irritability. These ventromedial and temporal subcomponents of the DMN may be crucial for integrating information about the self and environment and also the regulation of behavior and physiology (46).

Broadly, our findings align with a recent study demonstrating that youth-rated irritability can be predicted by brain networks during frustration (14). Prior work using the same task in independent samples also associates irritability with aberrant responses to or following FNR in FTL components (10,12). The FTL module (**Figure 4**) comprises fronto-limbic connections that are central to emotion regulation (47). Interestingly, frustration-increase was predicted by higher post-task E_*glob*_ in the FTL, while youth-rated irritability was predicted by lower FTL E_*glob*_. High E_*glob*_ is usually associated with better performance (31). A stronger increase in frustration usually elicits greater emotion regulation post-task, hence higher FTL E_*glob*_ was associated with frustration. However, this regulatory process might be impaired in irritability, which was therefore associated with lower FTL E_*glob*_. These hypotheses should be investigated further.

Discrepancies between parent and youth irritability-ratings are well-documented (48). Evidence suggests that each informant captures unique aspects of the phenotype (49) that are grounded in neurobiology (50). Here, parent-rated irritability was positively related to the efficiency of two post-task modules. Efficiency of vFSC that emerged after the task and included nodes such as ventral PFC and striatal regions, predicted parent-rated irritability, which has been previously associated with abrrant activity in these regions during frustration (11). While we found that efficiency of the post-task SMP module also predicted parent-rated irritability, FNR-responses of this network have been previously associated with youth-rated irritability (14), and structural abnormalities in motor circuits have been associated with both parent- and youth-irritability-ratings (51). Overall, our work supports the role of aberrant reward and motor circuitry in irritability, and highlights the need for research on mechanisms mediating informant effects.

## Limitations

A clear limitation of the present study is the small sample size, raising concerns regarding replicability. To mitigate this, we used regression models with a train/validation/test procedure instead of simple correlations for Aim 2. Nevertheless, replication in a larger sample is warranted. Also, we did not include a control session to rule out the possibility that pre- vs. post-RS differences relate to the attention task or the passage of time. Finally, we did not obtain frustration ratings after post-RS and, thus, cannot know whether the observed changes relate to the continued experience of frustration.

## Conclusions

We demonstrate that frustration induces more localized processing in the brain; a shift driven largely by reconfigurations of FP and aDMN-TL modules that persist into the post-frustration “recovery period.” The capacity for parallel information processing of modules that emerge during this recovery period explains individual differences in irritability and thus, may be a possible treatment target.

## Supporting information

Combined Supplement

## Data Availability

The data that support the findings of this study are available on request from the corresponding author (JOL). The data are not publicly available as not all participants consented to the sharing of their data.

## Acknowledgements

The authors’ research is supported by the National Institute of Mental Health (NIMH) Intramural Research Program (ZIAMH002786, ZIAMH002778, ZIAMH002782), conducted under NIH Clinical Study Protocols described at ClinicalTrials.gov (NCT02531893, NCT00025935, and NCT00018057).

## Author Contributions

Conceptualisation: JOL, AC, WLT, PS, MAB, SJG, EL, KK; Assessment and preprocessing: EX, LN, CZ, OR, SP, AJR; Supervision: JOL, AC, SPH, WLT, EL, KK; Formal analysis: JOL, SJG, KK; Writing – original draft preparation: JOL, EL, KK; Writing – review and editing: JOL, SPH, EX, LN, AC, CZ, OR, SP, AJR, WLT, PS, MAB, DSP, SJG, EL, KK; Visualisation JOL, EX, LN; Funding acquisition: EL, DSP.

