## Supplementary material for "Persistent frustration-induced reconfigurations of brain networks predict individual differences in irritability": Combined Supplement

### *Supplemental Information*

### Methods

#### *Sample*

Sixty-six youth participated in this study (33 females, mean age: 14.0 years, SD=2.76, min=9.3, max=20.9). Forty-eight participants met criteria for a mental disorder as established by a licensed clinician using the Kiddie Schedule for Affective Disorders and Schizophrenia (Kaufman et al., 1997). Participants were diagnosed with at least one of the following disorders: disruptive mood dysregulation disorder (DMDD, n=14) and oppositional defiant disorder (n=4), two diagnoses for which irritability is a diagnostic criterion (American Psychiatric Association, 2013); attention-deficit/hyperactivity disorder (ADHD, n=26); separation anxiety disorder (n=6); social phobia (n=9); and panic disorder (n=3), where irritability is also common (Eyre et al., 2019; Kircanski et al., 2017). Twenty-five participants were taking psychotropic medication (antidepressants: n=9; anticonvulsants: n=3; antipsychotics: n=3; stimulants: n=13; non-stimulant ADHD medication: n=3). For each participant, we calculated the composite measure of medication load (Sackeim, 2001), which we used as a nuisance variable in our analyses. Exclusion criteria included neurological disorders, autism spectrum disorders, psychosis, bipolar disorders, substance use, MRI contraindications, and full-scale IQ<70. The sample comprised participants with different racial and ethnical backgrounds (White, non-Hispanic: n=30; White, Hispanic: n=7; Black or African American, non-Hispanic: n=11; multiple racial identities, non-Hispanic: n=11; multiple racial identities, Hispanic: n=5; American Indian or Alaska native, non-Hispanic: n=1). The socioeconomic status (Hollingshead, 1975) of participants ranged between 20 and 120 (M=39, SD=24). Participants over age 18 and parents of minor participants gave written informed consent after receiving a complete description of the study; minors gave written assent. Procedures were approved by the Institutional Review Board of the National Institute of Mental Health.

#### *Assessment of symptom dimensions*

Irritability was assessed on the day of scanning with the Affective Reactivity Index (ARI), a 6-item questionnaire asking about temper outbursts and grouchy mood (Stringaris et al., 2012) retrospectively for the past week. Parent and youth completed the questionnaire separately, given well-documented informant discrepancy (Evans et al., 2020). On average the sample had mild-moderate levels of irritability (parent: M=3.6, SD=3.60; youth: M=2.8, SD=3.19) (Kircanski et al., 2017). Given the transdiagnostic sample, we also measured anxiety symptoms with the Screen for Child Anxiety Related Emotional Disorders (SCARED) (Birmaher et al., 1997), and inattention and hyperactivity, the two core domains of ADHD, with the respective subscales of the Conners III ADHD Rating Scales (Conners et al., 2011). The distribution of the sample on the different scales is shown in **Supplementary Figure S1**.

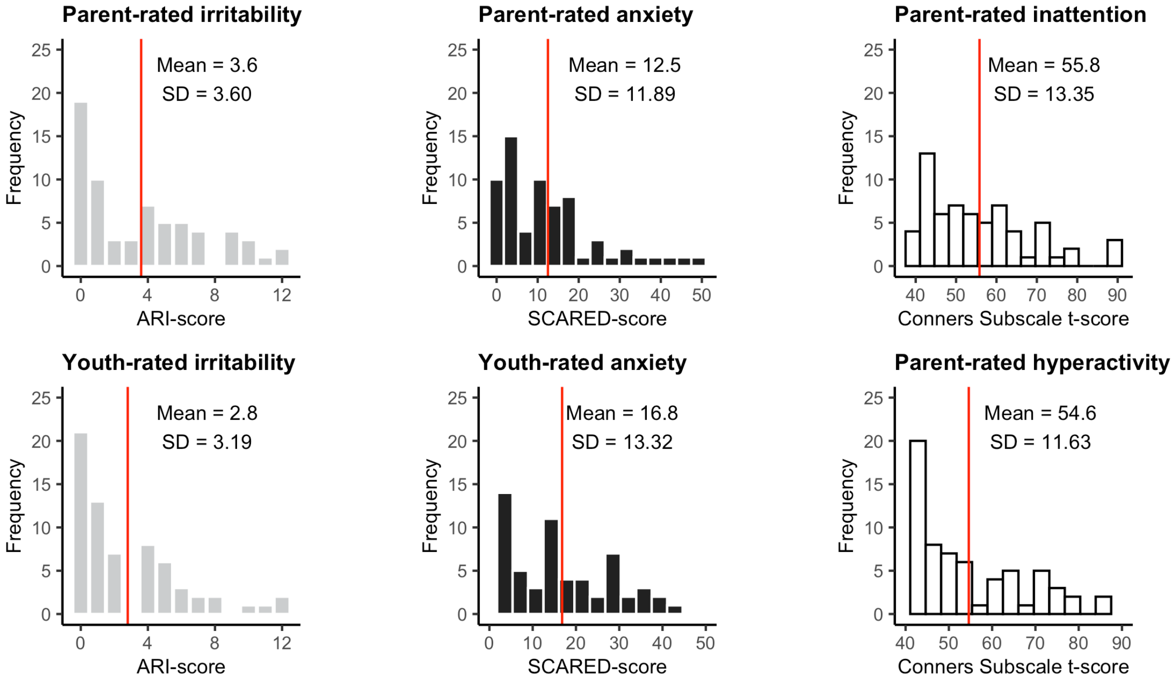


**Figure S1. Distribution of key symptom dimensions.** Irritability and anxiety were rated separately by parent and youth. Inattention and hyperactivity were rated by the parent only. The values for irritability and anxiety are the raw scores of the respective questionnaires.
**Abbreviations:** ***ARI,*** Affective Reactivity Index; ***Conners,*** Conners III ADHD Rating Scales; ***SCARED,*** Screen for Child Anxiety Related Emotional Disorders.

#### *Experimental Paradigm*

Participants completed a version of the attention-orienting Posner, which was adapted to induce frustration (**Figure 1**; Deveney et al., 2013; Tseng et al., 2019). The task itself is easy (~90% accuracy); following a cue that is a valid indicator of the target location in 75% of trials, participants indicate the target location (left or right side of the screen) via button press. Participants were instructed to respond as fast as possible and told that correct answers were followed by a monetary reward of $0.50.

First, a reward expectation was established during two 50-trial runs outside the scanner (Game 1). Next, participants entered the scanner and 9 minutes of baseline pre-task resting-state fMRI data were acquired. Then participants played the task again. However, unknown to participants, the contingencies were changed such that \$0.50 were deducted after a fixed proportion of correct responses, under the pretense that the response was "too slow." To ease participants into the frustrating task portion, they first completed 32 trials that contained 4 randomly spaced trials of rigged feedback while structural images were collected. Structural scanning was followed by two functional 50-trial frustration runs (Game 2). During these runs, rigged feedback was given for 60% of correct responses, causing the participants to lose a large proportion of their winnings from the preceding three runs. Feelings of frustration and unhappiness were assessed after each run using 9-point Likert scales (i.e., 1 = "happy" or "not at all frustrated"; 9 = "sad" or "extremely frustrated"). The task was followed by another 9 minutes of post-task resting-state data collection. Throughout the paradigm, finger photoplethysmography was recorded, which can be used to analyze pulse rate (PR) (Giardino et al., 2002). Increases in PR reflect physiological arousal (Pitson et al., 1994) and have been associated with feelings of anger and frustration (Zhang et al., 2016). However, increases in PR are not specific to anger/frustration and also occur in the context of other emotions (Siedlecka & Denson, 2019).

We were interested in delineating changes in brain network configuration from pre-task resting state to the task, specifically during four task events: (1) feedback indicating the deduction of \$0.50 (FNR), (2) anticipation of feedback in trials following FNR (FNR+1), (3) feedback indicating winning \$0.50 (Reward) and (4) anticipation of feedback in trials following reward (Reward+1). Similarly, we were interested in identifying changes from the task-based configurations to the post-task resting state. Finally, we were interested whether brain network reconfigurations relate to (a) task performance, (b) frustration, and (c) trait irritability. We used reaction time and error rates during valid trials, which are cognitively less demanding, and invalid trials, which are cognitively more demanding, as performance measures. Youth-ratings of frustration and PR were used to index frustration. Irritability was assessed on the day of scanning with the Affective Reactivity Index (Stringaris et al., 2012) for the week prior to scanning.

#### *Behavioral and physiological data*

Behavioral and physiological data were analyzed in R. We utilized a Game x Validity repeated-measures analysis of variance (ANOVA) as implemented in rstatix with age and medication load as covariates to investigate task performance in terms of reaction times and accuracy. To examine frustration ratings, we used a Game x Run ANOVA with the same covariates. These analyses were repeated with ARI-, SCARED- and Conners-scores as additional covariates to examine how symptoms affect task performance.

PR was analyzed with linear mixed effects analysis using lme4. As fixed effects, we entered time (rest, rest, task, task, task, task, rest, rest) and age. As random effects, we had intercepts for subjects, and by-subject random slopes for the effect of time. We first compared a linear vs. a quadratic effect of time. Then we created models that additionally contained either parent-rated ARI, youth-rated ARI, increase in frustration, and, as a test of specificity of the irritability-related measures, youth- or parent-rated anxiety. These variables were added as fixed effects, and random effects with by-subject random slopes. Visual inspection of residual plots did not reveal any obvious deviations from homoscedasticity or normality. P-values were obtained by likelihood ratio tests of the full model with the effect in question against the model without the effect in question.

*Acquisition of imaging data*

Imaging data were acquired on two identical 3.0 Tesla General Electric Signa scanners using a 32-channel head coil. Blood-oxygen-level-dependent (BOLD) changes during rest were measured for 9 minutes with a multi-echo planar imaging sequence (TR=2000 msec, TE1/2/3=14.8 / 28.4 / 42.0msec, flip angle 77°, matrix size 64×64, 34 axial interleaved slices, slice thickness 3.8mm, bandwidth = 7812.5 Hz/Pixel). The BOLD-response during the two 7-minute task runs was assessed with a different echo planar imaging sequence (TR=2300 msec, TE=30 msec, flip angle 70°, matrix size 96x96, 44 axial interleaved slices, slice thickness 3mm, bandwidth=5208.3 Hz/Pixel). In addition, a T1-weighted, magnetization-prepared, rapid-acquisition gradient echo (MPRAGE) sequence was acquired (TE=min full, TI=425, flip angle=7°, FoV=256x256x256, 1mm$^3$ voxels).

*Image processing*

Quality of the imaging data was assessed using MRIQC version 0.15.2 (Esteban et al., 2017). Of the originally 80 participants, 14 were excluded, due to the presence of severe motion-related artefacts (framewise displacement > 0.5mm/repetition time for more than 30% of the images). For the remaining 66 participants, we used the automated processing pipeline FMRIPREP (v20.0.5) for preprocessing (Esteban et al., 2019). The four initial volumes were discarded from all timeseries. First a reference image and mask were created based on the initial functional volumes. Then a series of spatial transformations was applied in one step including slice time correction using 3dTshift from AFNI, motion correction using MCFLIRT from FSL (Smith et al., 2004), and co-registration to the structural volume using boundary-based registration with 9 degrees of freedom (Greve & Fischl, 2009). Physiological noise regressors were extracted with

CompCor (Behzadi et al., 2007), and framewise displacement (Power et al., 2014) was calculated for each functional run using the implementation in Nipype. Finally, ICA-based Automatic Removal of Motion Artifacts (AROMA) was used to non-aggressively denoise the time courses (Pruim et al., 2015). We refrained from motion scrubbing as it has been repeatedly shown that AROMA performs at least as well in reducing motion-related artefacts (Ciric et al., 2017; Parkes et al., 2018), while allowing a constant number of timepoints across participants. For the multi-echo resting-state data, the Tedana T2* workflow (DuPre et al., 2019) was used to create an optimally weighted combination of the three echo times. Structural scans were subjected to FreeSurfer processing from FMRIPREP. Processed resting-state and task time series were resampled from voxel space to surface space. The shortest time series contained 236 time points.

*Network construction*

We used a 100-region parcellation scheme with cortical parcels previously assigned to known intrinsic functional networks (Schaefer et al., 2018; Yeo et al., 2011). We also used FreeSurfer segmentation to add the nucleus accumbens, caudate, pallidum, putamen, amygdala, hippocampus, thalamus and ventral diencephalon (separately for the left and right hemispheres). Thus, the functional connectivity network comprised 116 nodes. We regressed out from the time series motion parameters, white matter signal, cerebrospinal fluid signal, the first three principal components from aCompCor and first three cosine variables, framewise displacement, the spatial standard deviation of the temporal difference data, and the ICA-AROMA components classified as head motion; global signal was not regressed out.

For the task-based functional connectivity analyses, we used a regression approach (Cole et al., 2014; Hearne et al., 2017). For each of the 116 brain regions, task conditions convolved with a canonical hemodynamic response function were regressed from the time series to remove variance associated with task-related coactivation (Cole et al., 2014; Hearne et al., 2017). After accounting for the hemodynamic lag, we created a time series of interest specific to each task event (FNR, FNR+1, Reward, Reward+1) in each brain region by concatenating the residual time series associated with the relevant events within and across the two runs.

Functional connectivity during rest and task was quantified using Pearson correlations transformed for normality using Fisher's z'-transformation. This resulted in six connectivity matrices per subject: one for resting-state before task, four representing the different task events (FNR, FNR+1, Reward, Reward+1), and one for resting-state after the task.

*Aim 1a: Does brain network segregation differ among pre-task resting states, task events and post-task resting state?*

First, we focused on modularity, a metric that describes how well the brain can be subdivided in non-overlapping groups of nodes (i.e., modules); modules are characterized by a high number of intra-module connections and a low number of inter-module connections (Fortunato, 2010). The extent to which the brain exhibits such a modular structure can be quantified by the modularity index (Q) (Newman, 2006), which we estimated using the Louvain greedy algorithm (Blondel et al., 2008) implemented in the Brain Connectivity Toolbox (Rubinov & Sporns, 2010). Consistent with previous work (Alavash et al., 2019; Bassett et al., 2011; Hearne et al., 2017), we set the structural resolution parameter $\gamma$ (Fortunato & Barthelemy, 2007) to unity. Q-values closer to one indicate a network partition with many within- and few between-module connections, relative to an appropriate random network null model. Higher Q-values indicate a higher segregation of the brain associated with more localized information processing, while lower Q-values can be interpreted as more integrated information processing (**Figure S2**).

Given the stochastic initialization of the greedy optimization, the algorithm, which identifies the highest Q-value, was applied 1000 times for each experimental condition. The highest Q-value was then used to compare modularity indices among pre- and post-task resting state and all four task events. Additionally, we investigated how different graph density thresholds (i.e., 5%, 10%, 15%, 20%, 25% or 30%) affected the results. Using the Permutation Analysis of Linear Models (PALM) (Winkler et al., 2016) and 5000 permutations, we compared Q-values between conditions using paired t-tests. Significant results were those that passed a threshold of $p < .05$ using family-wise error rate (FWER) correction across the six density thresholds. We used Hedges's *g* to calculate effect sizes.

### Illustration of expected changes in modularity

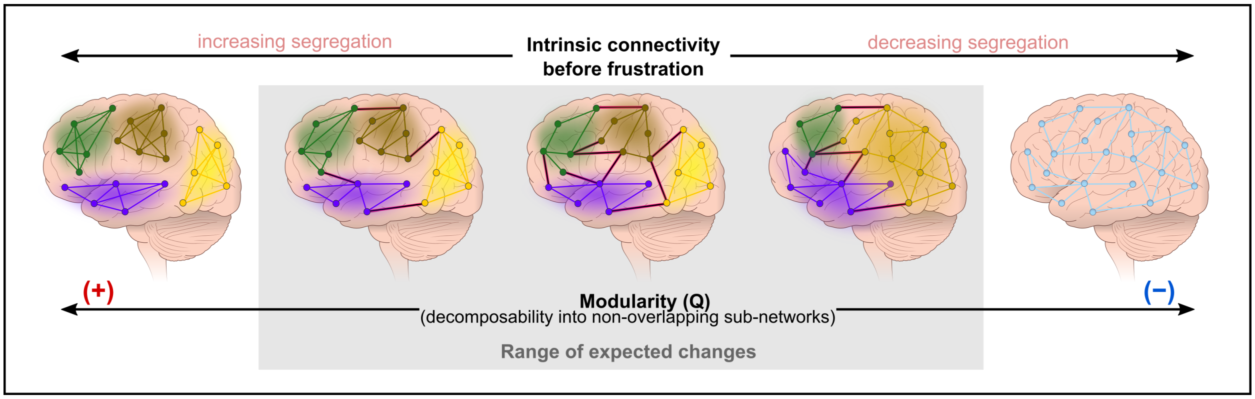


**Figure S2. Range of potential changes in the modular structure of the brain during frustration.** Depending on environmental events or internal states, the brain can transition to a more localized, segregated processing mode (left side of the figure) characterized by clearly separable sub-networks with many within, and few between, subnetwork connections. Alternatively, the brain can transition to a more integrated processing mode (right side of the figure).

*Aim 1b: Does the modular composition of brain networks differ among pre-task resting states, task events and post-task resting state?*

Because multiple module partitions maximize Q, we used a consensus approach to calculate an agreement matrix across the 1000 iterations (see above) for each participant and condition. These agreement matrices represent the tendency for each pair of nodes to be assigned to the same module across iterations and served as input to an independent module partitioning. Thus, we obtained one module partition for each participant and condition. In this step, we entered the original network structure from the parcellation scheme (Schaefer et al., 2018), used to extract the time courses, as a prior, and set the resolution parameter $\tau$ to 0.75.

Next, a group-level modularity partition was obtained. For each condition, we calculated a matrix reflecting the probability of nodes being assigned to the same module across all 66 participants (Hearne et al., 2017). These matrices were then subjected to the graph-theoretical community detection algorithm used at the individual level. The result of this analytic step is used for visualization of the changes in module composition across the experimental conditions.

Consistent with prior work (Dwyer et al., 2014; Hearne et al., 2017), we compared the nodal composition of modules across conditions with the variation of information metric (VIn) (Meilă, 2007), which quantifies the distance between modularity partitions. Statistically significant differences in modular structure were determined using a repeated-measures permutation procedure (Dwyer et al., 2014; Hearne et al., 2017). For half of the participants, the condition labels were randomly switched between experimental conditions (e.g., pre- vs. post-task resting-state) resulting in two shuffled sets of individual-level module structures. These were submitted to the original pipeline, which generated two new group-level module partitions for which we calculated the VIn. We repeated these steps 1000 times, which created a null distribution to which the original VIn-values were compared for each condition across network density levels.

If significant differences between two conditions were observed, we calculated the VIn for each module for each condition and participant to determine the relative contribution of a specific module to the reconfiguration (Braun et al., 2015; Hearne et al., 2017). To achieve this goal VIn values of the single modules were compared across the six network density thresholds using paired t-tests in PALM with 5000 permutations. A significant higher VIn value indicates a greater contribution of that network to the overall reconfiguration.

*Aim 2: In which subnetworks, measured at what phase of the paradigm (i.e., before, during or after the frustrating task), does information processing efficiency predict frustration and irritability?*

Lastly, we tested whether modules specific to either the resting-state conditions or task events were related to differences in pre- vs. post-task frustration-ratings or to trait irritability. To assess the impact of altered functional connectivity described in Aim 1 on behavioral measures, we calculated global efficiency ($E_{glob}$), which is defined as the inverse of the average path length between all nodes in a network (Latora & Marchiori, 2001). If we assume that information follows the shortest path, $E_{glob}$ quantifies the capacity for parallel information processing in a network. To date, numerous studies have linked $E_{glob}$ to neurophysiological (Cocchi et al., 2017; de Pasquale et al., 2016), cognitive (Bassett et al., 2009; Hearne et al., 2017) and emotional processes (Pan et al., 2018). So, for each participant, $E_{glob}$ was calculated within each module from the group-level modularity partition at the fixed network density of 10%. To determine the replicability of our findings, we used a prediction framework, rather than simple correlations, to identify modules relevant to increases in frustration ratings and interindividual differences in irritability. We did this by dividing our sample into training/validation and held-out datasets (80/20). We used stratified random sampling to ensure that both datasets contained an equal percentage of diagnostic categories (healthy volunteers; individuals with diagnoses of ADHD, DMDD or an anxiety disorder) had an equal mean age and sex ratio. We used a linear stepwise regression (leapSeq), applying a 10-fold cross validation with 20 repeats as implemented in the Caret package for R, to select predictors of frustration increase and irritability. In addition to the efficiency of all brain modules during the two resting state conditions and the four task events, predictors in the model included age (previously associated with individual differences in irritability (Copeland et al.,

2015), sex, medication load (Sackeim, 2001), and scanner. The identified model was then used to predict change in frustration ratings, youth- and parent-rated irritability in the held-out dataset. We used 5000 permutations to test whether the association between the predicted and real values in the held-out set were significant at a threshold of $p_{FDR} < .05$ applying False Discovery Rate correction across the three models. Given the diagnostic status of the sample, we also tested models designed to predict ratings of anxiety, inattention and hyperactivity.

### Behavioral Results

Overall, participants responded faster ($F_{(1,63)}$=20.66, p<.001, $\eta^2$=.25) and more accurately ($F_{(1,63)}$=21.85, p<.001, $\eta^2$=.27) during valid compared to invalid trials. This effect was stronger during Game 2 that contained the frustrating condition (Game×Validity, reaction time: $F_{(1,63)}$=9.60, p=.003, $\eta^2$=.13, accuracy: $F_{(1,63)}$=7.95, p=.006, $\eta^2$=.11). During the invalid condition, older participants responded faster (Validity×Age: $F_{(1,63)}$=6.06, p=.017, $\eta^2$=.09) and more accurately (Validity×Age: $F_{(1,63)}$=5.82, p<.019, $\eta^2$=.09). Higher levels of attention problems predicted lower accuracy in invalid trials (Validity×Inattention: $F_{(1,63)}$=5.12, p=.027, $\eta^2$=.08). Reaction times and accuracy did not depend on medication load, levels of irritability, anxiety or hyperactivity (all p-values >.082).

The introduction of the rigged feedback was associated with increased frustration ($F_{(1,63)}$=18.26, p<.001, $\eta^2$=.23) and sadness ratings ($F_{(1,63)}$=7.45, p=.008, $\eta^2$=.11). Increased frustration was more pronounced in younger participants (Game×Age: $F_{(1,63)}$=5.27, p=.025, $\eta^2$=.08), and participants that reported higher levels of anxiety (Game×SCARED-Y: $F_{(1,59)}$=6.28, p=.015, $\eta^2$=.10). Age (Game×Age: $F_{(1,63)}$=2.17, p=.146, $\eta^2$=.03) and anxiety (Game×Age: $F_{(1,59)}$=0.83, p=.366, $\eta^2$=.01) did not relate to the increase in sadness. There was no effect of medication load, symptoms of inattention or hyperactivity, parent-reported levels of anxiety or parent or youth-reported irritability on the increase in frustration or sadness ratings (all p-values >.089).

Modeling PR as a quadratic function of time fit the data significantly better than a model using a linear function ($\chi^2_{(4)}=1.55\times10^{-5}$). The model fit improved when age ($\beta$=−3.52, $t_{(56.02)}$=−2.64, p=.011) was added as a fixed effect ($\chi^2_{(1)}$=.015). Specifically, elevated PR was seen in younger participants after the task (all p-values<.025, all Hedges' g>.70). Fit between model and data further improved when parent-ratings of irritability ($\beta$=3.66, $t_{(56.00)}$=2.83, p=.007) were added as fixed effect ($\chi^2_{(1)}$=.011); participants with higher parent-rated irritability during and after the task (all p-values<.037, all Hedges' g>.65; **Figure 1C**). Youth-ratings of irritability ($\beta$=1.45, $t_{(56.20)}$=1.07, p=.288), increase in frustration ratings ($\beta$=1.94, $t_{(55.99)}$=1.36, p=.181), parent-rated anxiety ($\beta$=1.83, $t_{(56.25)}$=1.29, p=.202) and youth-rated anxiety ($\beta$=1.99, $t_{(56.06)}$=1.67, p=.100) were not predictors of PR changes throughout the paradigm.

### Effects of graph thresholding on one-by-one modularity findings

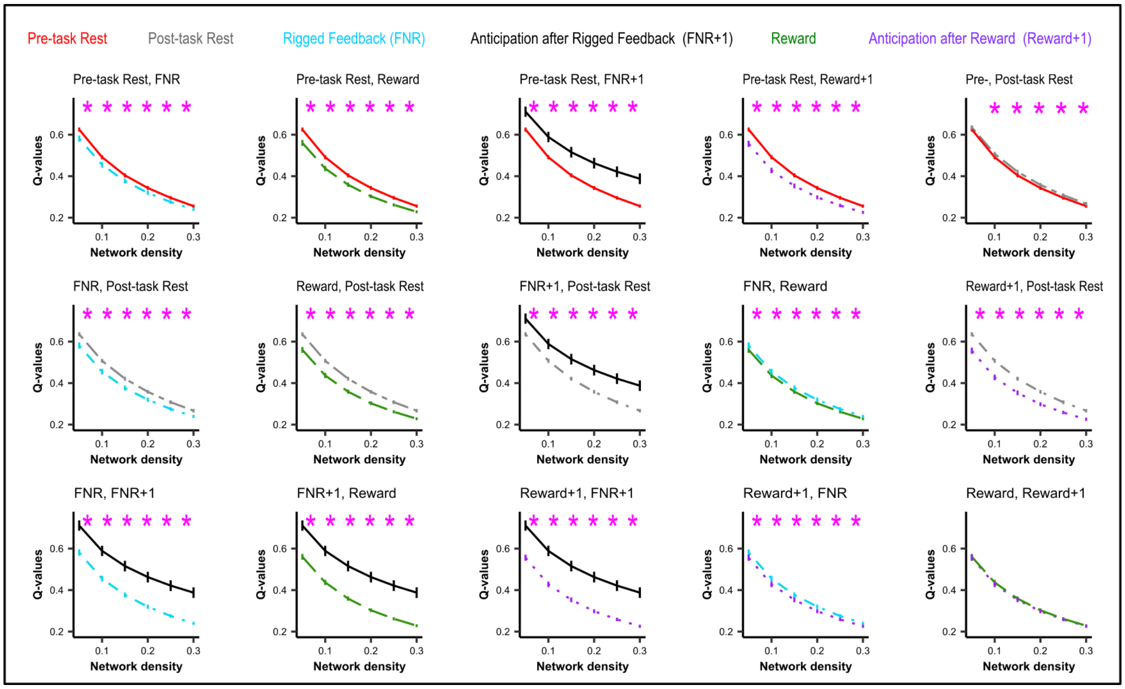


**Figure S3. One-by-one comparison of the four task and two resting state conditions across a range of thresholds (5 – 30%).** Pink stars indicate significance of the respective pair-wise comparison at $p_{FWE} < .05$. This underscores that differences between task conditions reported in the main manuscript are not specific to a certain network density. Error bars represent standard error of the mean.

### Overlap of the nodal network affiliations between the Schaefer atlas and the pre-task resting state data

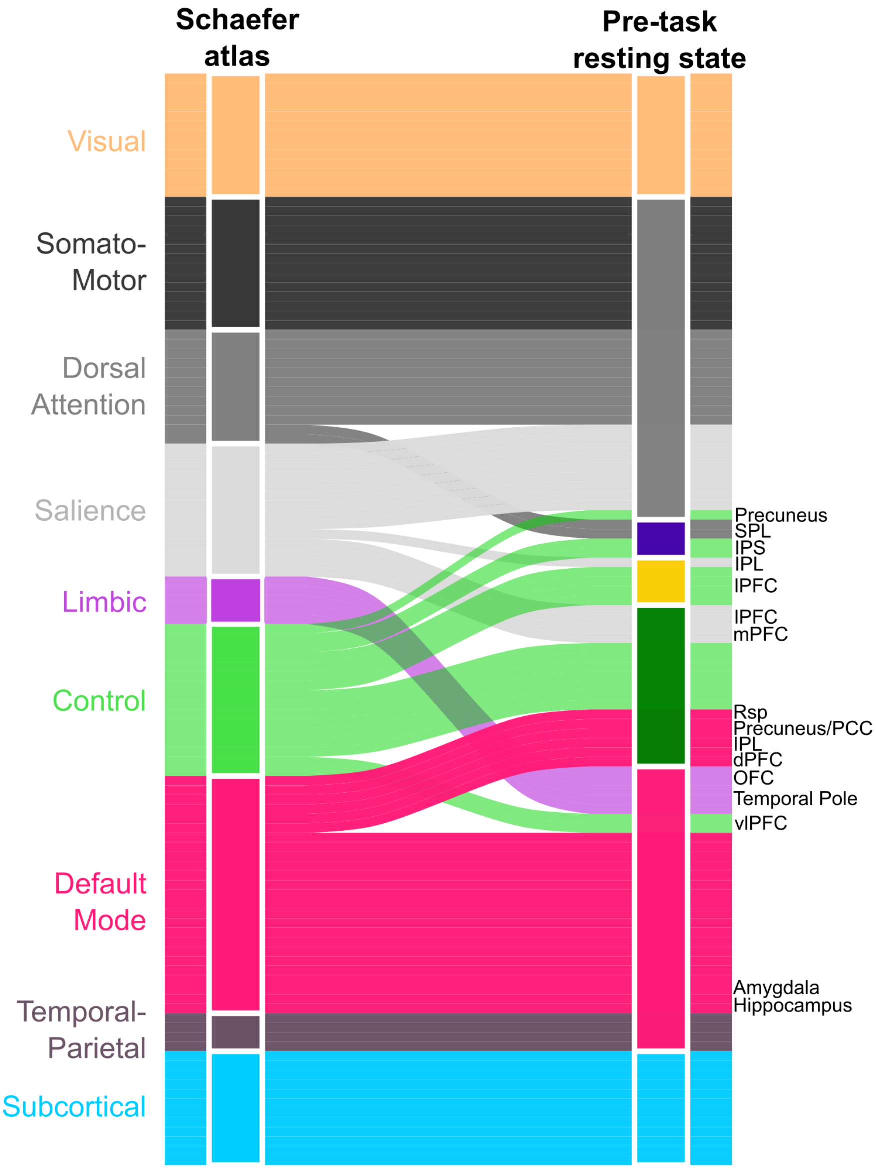


**Figure S4. Nodal network affiliations within the Schaefer atlas and our pre-task resting state data.** Network names at the left are taken from the Schaefer atlas. The subcortical network (light blue) was not part of the Schaefer atlas, but comprises the subcortical regions taken from the FreeSurfer segmentation. In our resting state data, the somato-motor, dorsal attention and salience network are merged to represent one large network. The anterior default mode, temporal and limbic networks are also merged in our data. The control network, however, branches into three different networks in our data. At the right side, we show the names of the nodes that are assigned to a different network in our data vs. the atlas (Schaefer et al., 2018). Further, we highlight that the amygdala and hippocampus, not contained in the Schaefer atlas, are assigned to the anterior default mode-temporal-limbic module in our data.

**Table S1. Variation of information (VIn) statistics for the six experimental conditions across levels of network density.**

| | Network density | | | | | | | | | | | |
|---|---|---|---|---|---|---|---|---|---|---|---|---|
| | 5% | | 10% | | 15% | | 20% | | 25% | | 30% | |
| | VIn | *p* | VIn | *p* | VIn | *p* | VIn | *p* | VIn | *p* | VIn | *p* |
| **Pre-RS** | | | | | | | | | | | | |
| Reward | .131 | 1 | .155 | .009 | .188 | .014 | .203 | .162 | .257 | <.001 | .261 | .194 |
| Reward+1 | .189 | .064 | .300 | <.001 | .284 | <.001 | .290 | <.001 | .268 | <.001 | .293 | .002 |
| FNR | .175 | .869 | .115 | .777 | .115 | .677 | .122 | .809 | .195 | .313 | .212 | .822 |
| FNR+1 | .162 | .973 | .176 | <.001 | .206 | .004 | .248 | .004 | .250 | <.001 | .291 | .002 |
| Post-RS | .134 | 1 | .223 | <.001 | .197 | .001 | .197 | .006 | .203 | .006 | .262 | .002 |
| **Reward** | | | | | | | | | | | | |
| Reward+1 | .142 | 1 | .240 | <.001 | .181 | .341 | .191 | .319 | .164 | 1 | .121 | 1 |
| FNR | .138 | 1 | .110 | .918 | .111 | 1 | .144 | .797 | .133 | 1 | .156 | 1 |
| FNR+1 | .110 | 1 | .122 | .061 | .172 | .281 | .112 | .990 | .158 | 1 | .143 | 1 |
| Post-RS | .187 | 1 | .260 | .005 | .286 | <.032 | .293 | <.001 | .329 | <.001 | .322 | <.001 |
| **Reward+1** | | | | | | | | | | | | |
| FNR | .146 | .209 | .232 | <.001 | .241 | <.001 | .192 | .062 | .194 | .464 | .179 | .515 |
| FNR+1 | .126 | 1 | .202 | .004 | .183 | .763 | .140 | 1 | .129 | 1 | .105 | 1 |
| Post-RS | .277 | <.001 | .358 | <.001 | .336 | <.001 | .336 | <.001 | .349 | <.001 | .327 | <.001 |
| **FNR** | | | | | | | | | | | | |
| FNR+1 | .173 | .007 | .103 | .002 | .174 | .005 | .164 | .003 | .151 | .006 | .164 | .009 |
| Post-RS | .222 | .838 | .260 | .525 | .216 | .817 | .207 | .749 | .224 | .603 | .295 | <.001 |
| **FNR+1** | | | | | | | | | | | | |
| Post-RS | .241 | .196 | .232 | .960 | .223 | 0.397 | .256 | .270 | .285 | .042 | .244 | .565 |

**Abbreviations:** ***p***, p-value; ***Reward+1***, Anticipation following reward; ***FNR***, Frustrative non-reward operationalized as rigged feedback; ***FNR+1***, Anticipation following rigged feedback; ***VIn***, variation of information

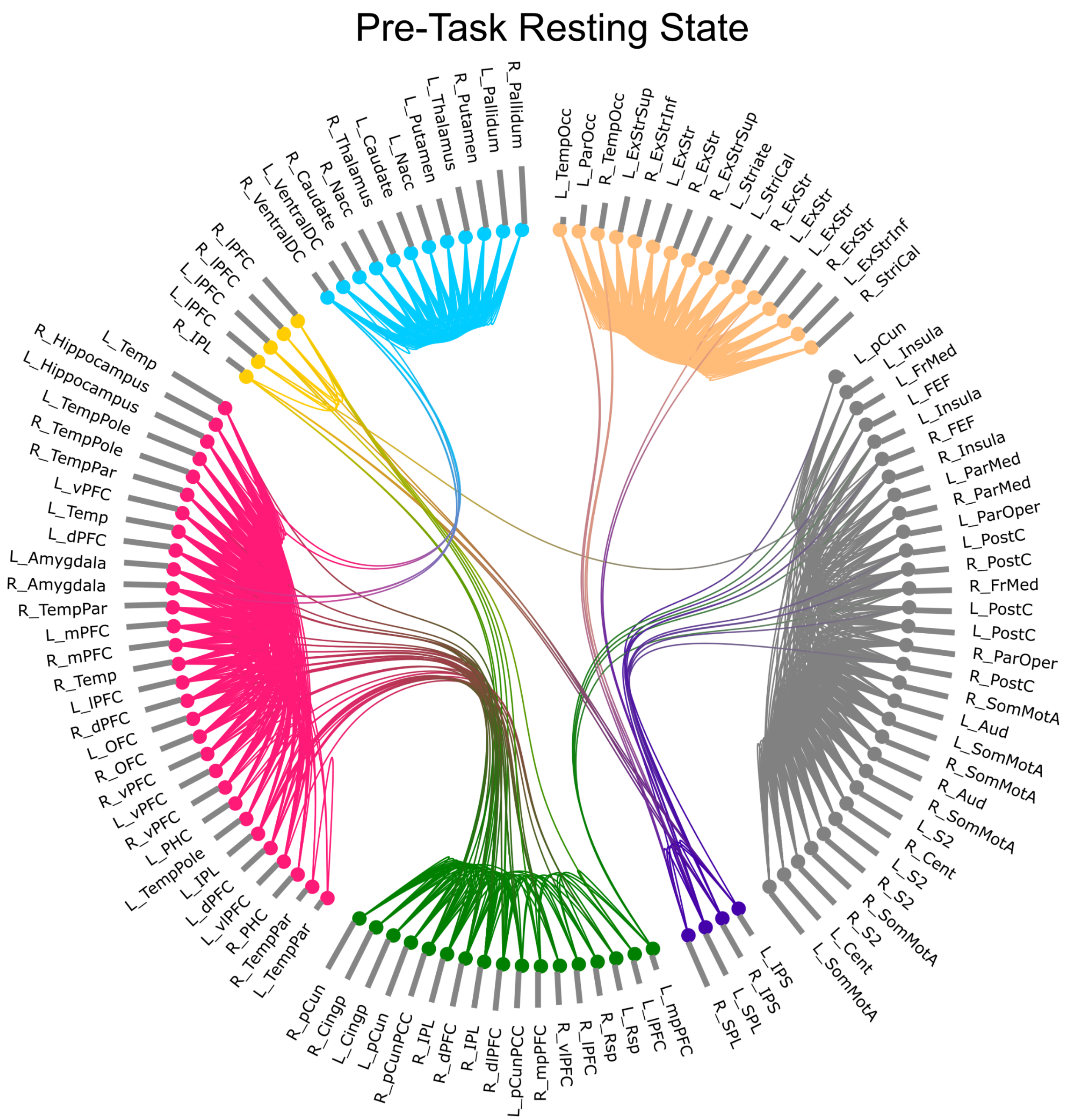


**Figure S5. Modular composition and nodal network during pre-task resting state.** The color scheme corresponds to Figure 3A in the main manuscript. Within each module nodes are sorted by centrality (represented by the bar attached to each node) within the specific module. We show the top 15% of connections as lines between the nodes. For an interactive version of this figure please see: https://immersive.erc.monash.edu/neuromarvl/?save=8d9ec8a0-a088-495e-86a4-196ca85326c9_128.231.234.5

**Abbreviations:** ***dPFC***, dorsal prefrontal cortex; ***FEF,*** frontal eye field; ***FrMed,*** medial frontal cortex; ***IPL,*** inferior parietal lobe; ***IPS,*** intraparietal sulcus; ***L***, left; ***mPFC,*** medial prefrontal cortex; ***mPFC***, medial prefrontal cortex; ***Nacc,*** nucleus accumbens; ***OFC,*** orbitofrontal cortex; ***ParMed,*** medial parietal cortex; ***ParOper,*** parietal operculum; ***PHC***, parahippocampal cortex; ***R***, right; ***SomMot,*** somato-motor cortex; ***SPL,*** superior parietal lobe; ***Temp***, temporal; ***TempPar,*** temporoparietal region; ***TempPole***, temporal pole; ***ventral DC,*** ventral diencephalon, which includes hypothalamus, mammillary bodies, subthalamic nuclei, substantia nigra, red nucleus, and medial and lateral geniculate nuclei; ***vPFC***, ventro-medial prefrontal cortex; ***vlPFC***, ventro-lateral prefrontal cortex

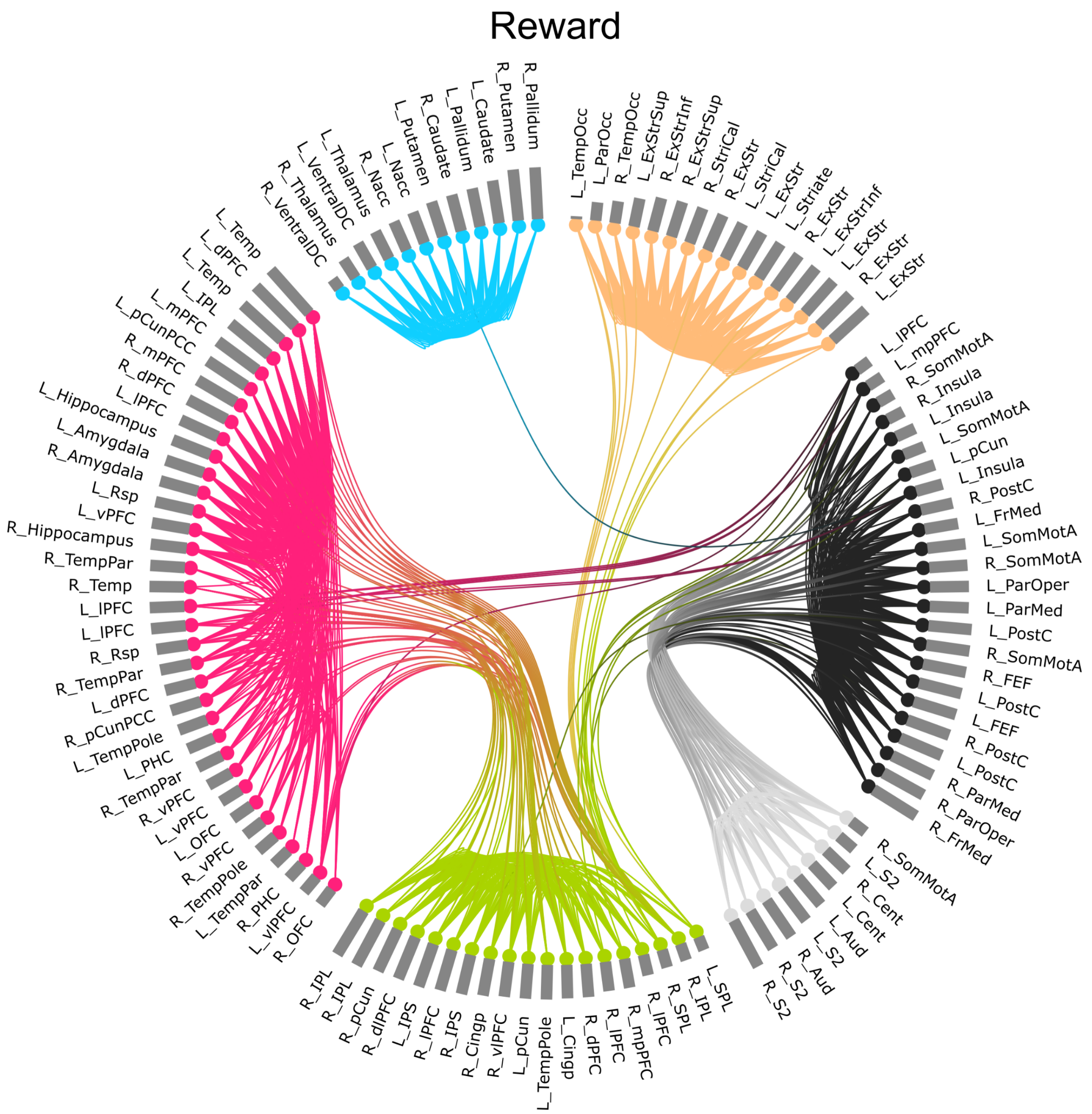


**Figure S6. Modular composition and nodal network during Reward.** The color scheme corresponds to Figure 3A in the main manuscript. Within each module nodes are sorted by centrality (represented by the bar attached to each node) within the specific module. We show the top 15% of connections as lines between the nodes. For an interactive version of this figure please see: https://immersive.erc.monash.edu/neuromarvl/?save=4afb3247-c7b2-4284-9348-8294736d4124_128.231.234.7

**Abbreviations:** ***dPFC***, dorsal prefrontal cortex; ***FEF,*** frontal eye field; ***FrMed,*** medial frontal cortex; ***IPL,*** inferior parietal lobe; ***IPS,*** intraparietal sulcus; ***L***, left; ***mPFC,*** medial prefrontal cortex; ***mPFC***, medial prefrontal cortex; ***Nacc,*** nucleus accumbens; ***OFC,*** orbitofrontal cortex; ***ParMed,*** medial parietal cortex; ***ParOper,*** parietal operculum; ***PHC***, parahippocampal cortex; ***R***, right; ***SomMot,*** somato-motor cortex; ***SPL,*** superior parietal lobe; ***Temp***, temporal; ***TempPar,*** temporoparietal region; ***TempPole***, temporal pole; ***ventral DC,*** ventral diencephalon, which includes hypothalamus, mammillary bodies, subthalamic nuclei, substantia nigra, red nucleus, and medial and lateral geniculate nuclei; ***vPFC***, ventro-medial prefrontal cortex; ***vlPFC***, ventro-lateral prefrontal cortex

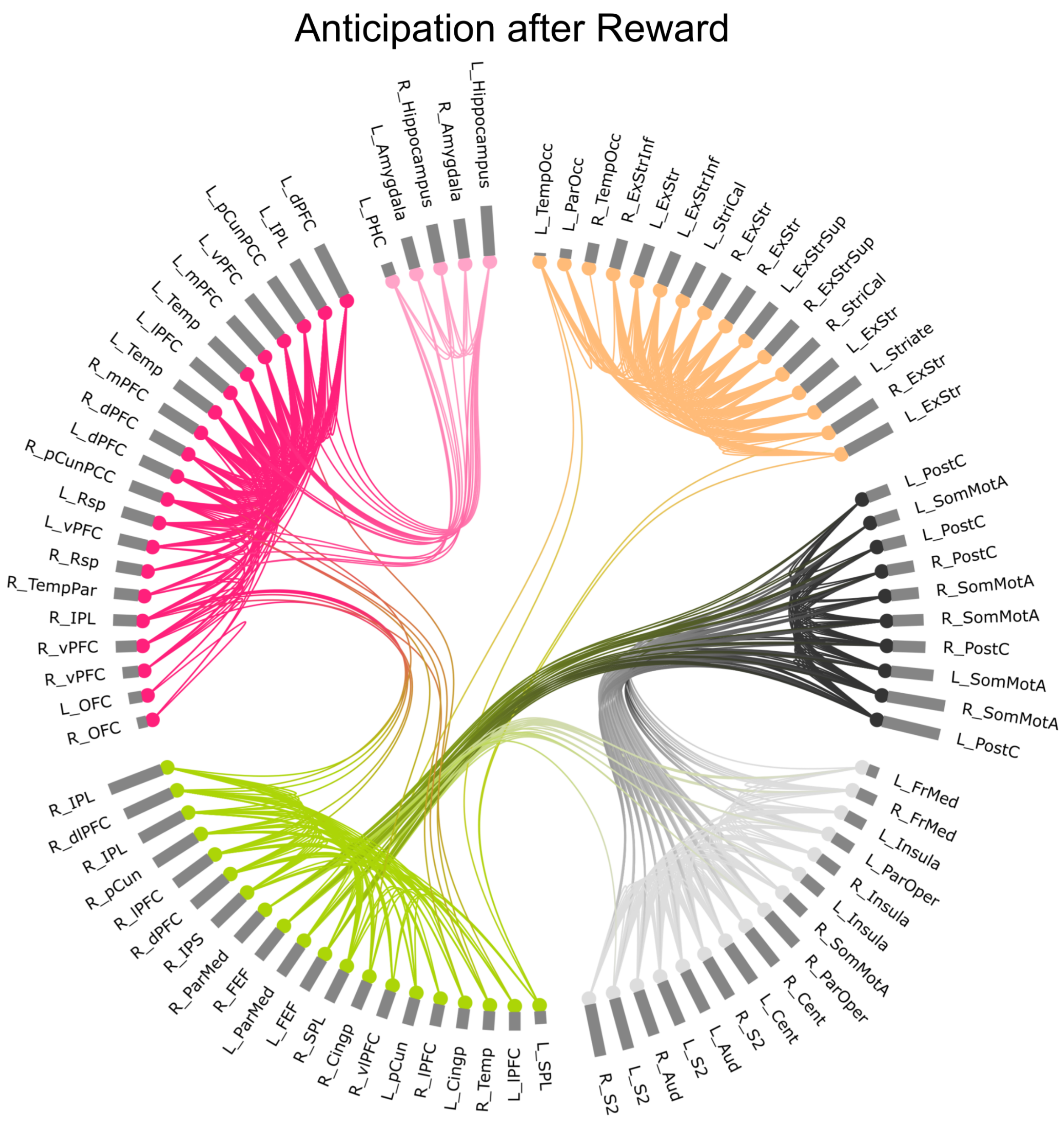


**Figure S7. Modular composition and nodal network during anticipation following reward (Reward+1).** The color scheme corresponds to Figure 3A in the main manuscript. Within each module nodes are sorted by centrality (represented by the bar attached to each node) within the specific module. We show the top 15% of connections as lines between the nodes. For an interactive version of this figure please see: https://immersive.erc.monash.edu/neuromarvl/?save=db79a3e2-d70e-4f83-a09c-30a143ddb02e_128.231.234.5

**Abbreviations:** ***dPFC***, dorsal prefrontal cortex; ***FEF,*** frontal eye field; ***FrMed,*** medial frontal cortex; ***IPL,*** inferior parietal lobe; ***IPS,*** intraparietal sulcus; ***L***, left; ***mPFC,*** medial prefrontal cortex; ***mPFC***, medial prefrontal cortex; ***Nacc,*** nucleus accumbens; ***OFC,*** orbitofrontal cortex; ***ParMed,*** medial parietal cortex; ***ParOper,*** parietal operculum; ***PHC***, parahippocampal cortex; ***R***, right; ***SomMot,*** somato-motor cortex; ***SPL,*** superior parietal lobe; ***Temp***, temporal; ***TempPar,*** temporoparietal region; ***TempPole***, temporal pole; ***ventral DC,*** ventral diencephalon, which includes hypothalamus, mammillary bodies, subthalamic nuclei, substantia nigra, red nucleus, and medial and lateral geniculate nuclei; ***vPFC***, ventro-medial prefrontal cortex; ***vlPFC***, ventro-lateral prefrontal cortex

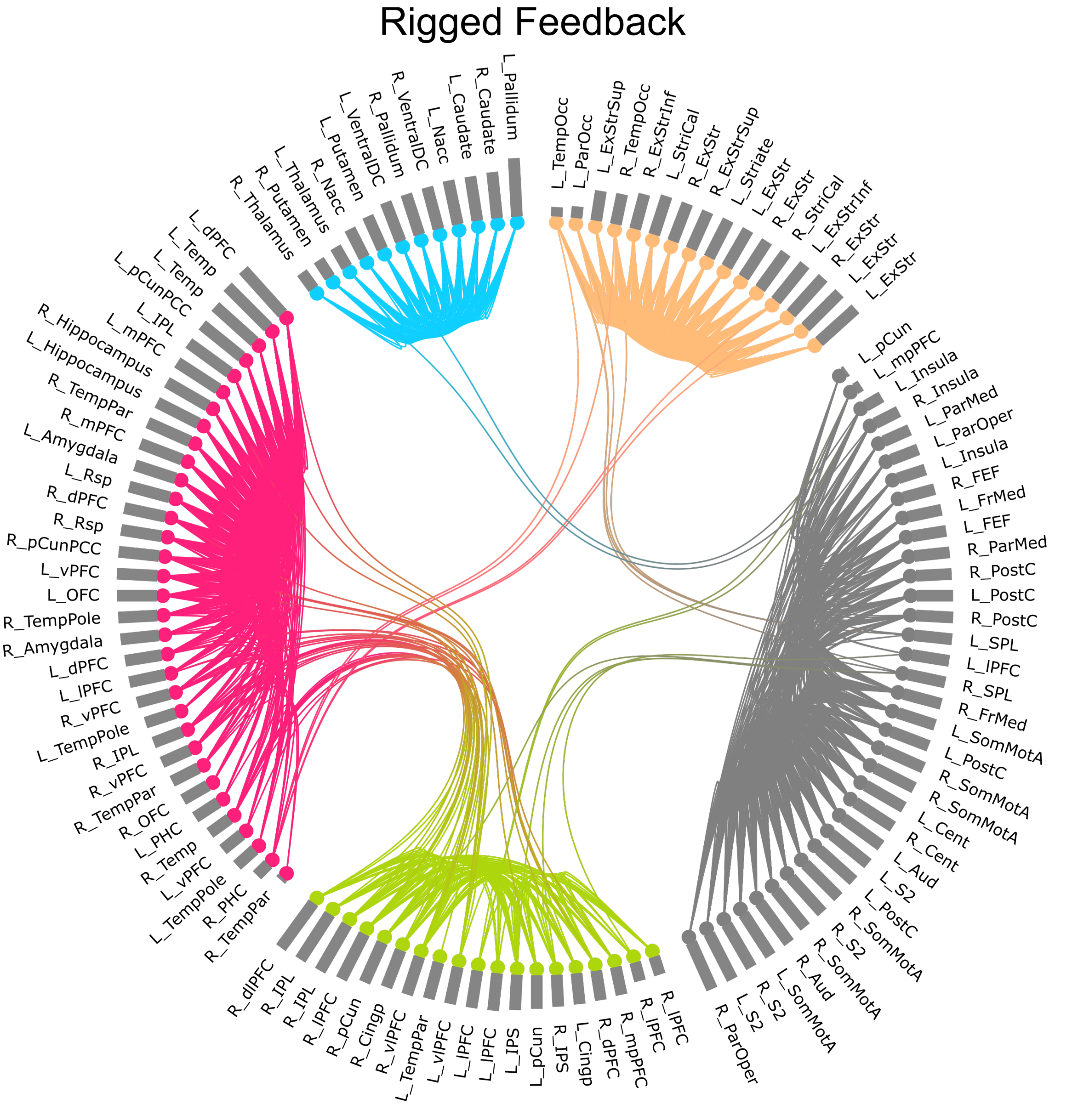


**Figure S8. Modular composition and nodal network during rigged feedback (FNR).** The color scheme corresponds to Figure 3A in the main manuscript. Within each module nodes are sorted by centrality (represented by the bar attached to each node) within the specific module. We show the top 15% of connections as lines between the nodes. For an interactive version of this figure please see: https://immersive.erc.monash.edu/neuromarvl/?save=e0804aa3-2026-4e10-9eb7-83fe14a76cf5_128.231.2.239

**Abbreviations:** ***dPFC***, dorsal prefrontal cortex; ***FEF,*** frontal eye field; ***FrMed,*** medial frontal cortex; ***IPL,*** inferior parietal lobe; ***IPS,*** intraparietal sulcus; ***L***, left; ***mPFC,*** medial prefrontal cortex; ***mPFC***, medial prefrontal cortex; ***Nacc,*** nucleus accumbens; ***OFC,*** orbitofrontal cortex; ***ParMed,*** medial parietal cortex; ***ParOper,*** parietal operculum; ***PHC***, parahippocampal cortex; ***R***, right; ***SomMot,*** somato-motor cortex; ***SPL,*** superior parietal lobe; ***Temp***, temporal; ***TempPar,*** temporoparietal region; ***TempPole***, temporal pole; ***ventral DC,*** ventral diencephalon, which includes hypothalamus, mammillary bodies, subthalamic nuclei, substantia nigra, red nucleus, and medial and lateral geniculate nuclei; ***vPFC***, ventro-medial prefrontal cortex; ***vlPFC***, ventro-lateral prefrontal cortex

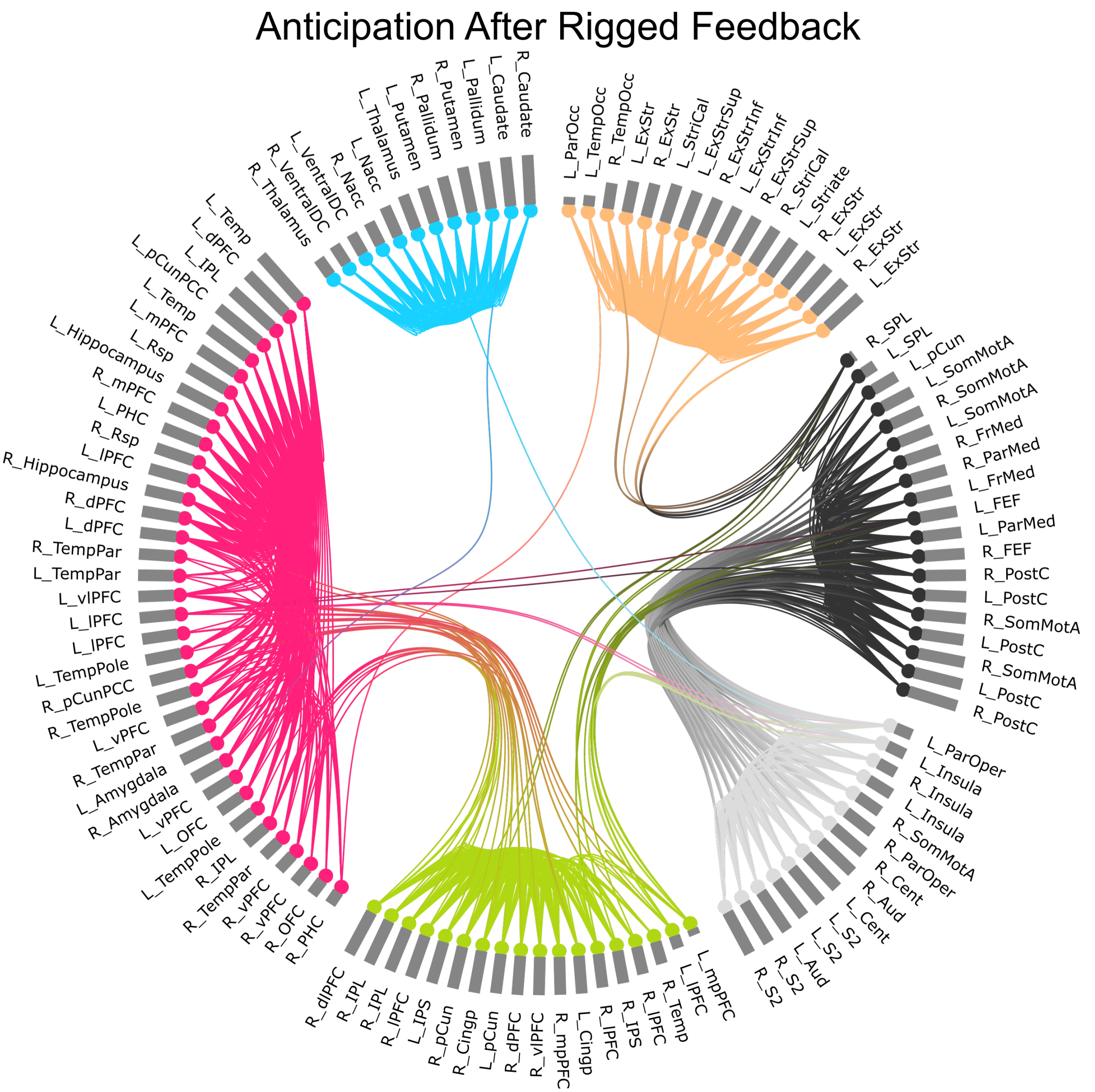


**Figure S9. Modular composition and nodal network during anticipation following rigged feedback (FNR+1).** The color scheme corresponds to Figure 3A in the main manuscript. Within each module nodes are sorted by centrality (represented by the bar attached to each node) within the specific module. We show the top 15% of connections as lines between the nodes. For an interactive version of this figure please see: https://immersive.erc.monash.edu/neuromarvl/?save=15840252-c739-49f6-bb80-c608c41e04c3_128.231.2.239

**Abbreviations:** ***dPFC***, dorsal prefrontal cortex; ***FEF,*** frontal eye field; ***FrMed,*** medial frontal cortex; ***IPL,*** inferior parietal lobe; ***IPS,*** intraparietal sulcus; ***L***, left; ***mPFC,*** medial prefrontal cortex; ***mPFC***, medial prefrontal cortex; ***Nacc,*** nucleus accumbens; ***OFC,*** orbitofrontal cortex; ***ParMed,*** medial parietal cortex; ***ParOper,*** parietal operculum; ***PHC***, parahippocampal cortex; ***R***, right; ***SomMot,*** somato-motor cortex; ***SPL,*** superior parietal lobe; ***Temp***, temporal; ***TempPar,*** temporoparietal region; ***TempPole***, temporal pole; ***ventral DC,*** ventral diencephalon, which includes hypothalamus, mammillary bodies, subthalamic nuclei, substantia nigra, red nucleus, and medial and lateral geniculate nuclei; ***vPFC***, ventro-medial prefrontal cortex; ***vlPFC***, ventro-lateral prefrontal cortex

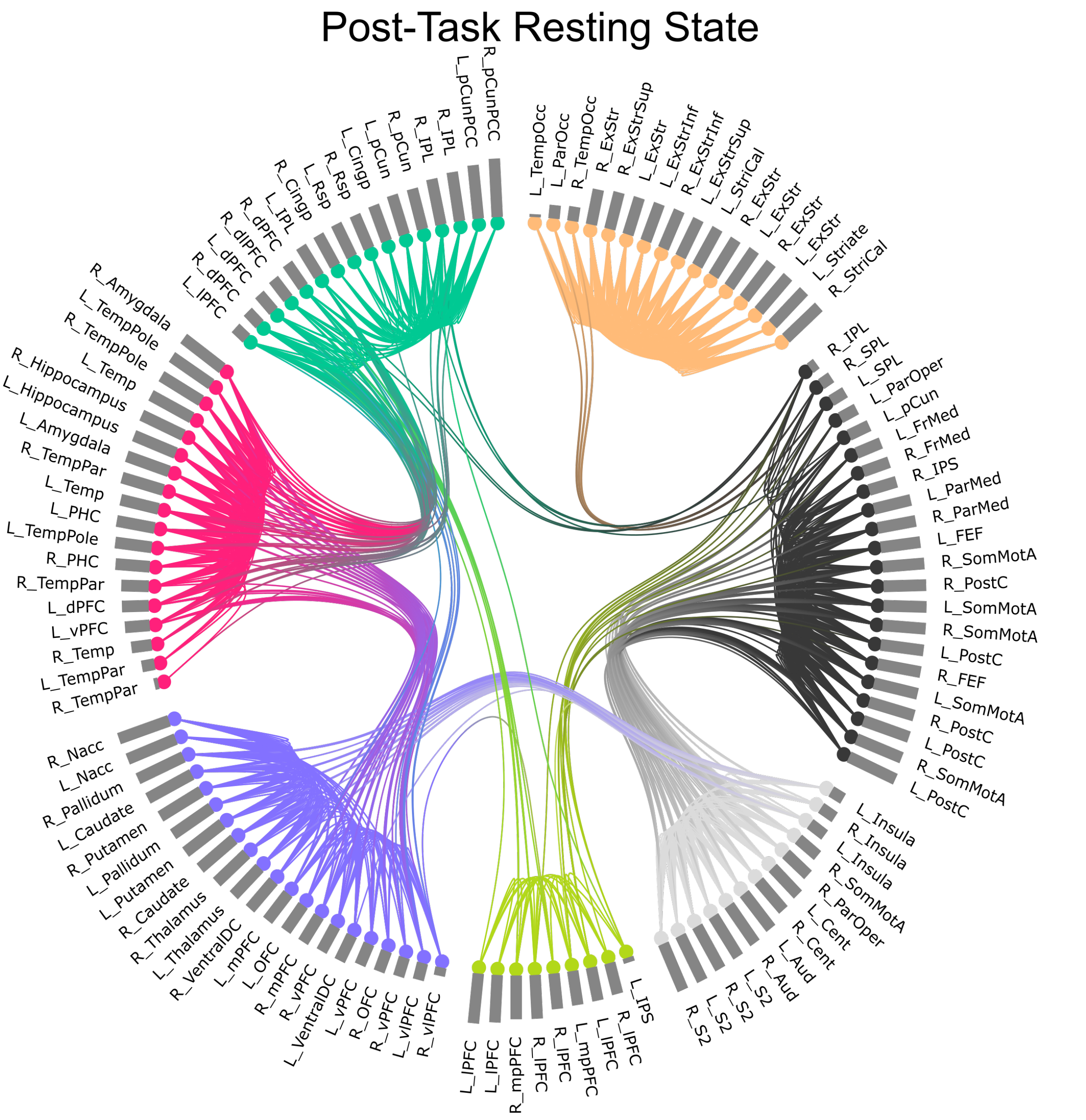


**Figure S10. Modular composition and nodal network during post-task resting state.** The color scheme corresponds to Figure 3A in the main manuscript. Within each module nodes are sorted by centrality (represented by the bar attached to each node) within the specific module. We show the top 15% of connections as lines between the nodes. For an interactive version of this figure please see: https://immersive.erc.monash.edu/neuromarvl/?save=a00fbada-5268-4088-8974-9d1622b70f9a_128.231.2.239

**Abbreviations:** ***dPFC***, dorsal prefrontal cortex; ***FEF,*** frontal eye field; ***FrMed,*** medial frontal cortex; ***IPL,*** inferior parietal lobe; ***IPS,*** intraparietal sulcus; ***L***, left; ***mPFC,*** medial prefrontal cortex; ***mPFC***, medial prefrontal cortex; ***Nacc,*** nucleus accumbens; ***OFC,*** orbitofrontal cortex; ***ParMed,*** medial parietal cortex; ***ParOper,*** parietal operculum; ***PHC***, parahippocampal cortex; ***R***, right; ***SomMot,*** somato-motor cortex; ***SPL,*** superior parietal lobe; ***Temp***, temporal; ***TempPar,*** temporoparietal region; ***TempPole***, temporal pole; ***ventral DC,*** ventral diencephalon, which includes hypothalamus, mammillary bodies, subthalamic nuclei, substantia nigra, red nucleus, and medial and lateral geniculate nuclei; ***vPFC***, ventro-medial prefrontal cortex; ***vlPFC***, ventro-lateral prefrontal cortex

### Additional Analyses to rule out contamination by motion

Our first set of analyses focused on modularity. To ensure that findings were not attributable to motion, we calculated the mean framewise displacement for each of the six conditions (pre-task resting state, Reward, Reward+1, FNR, FNR+1, post-task resting state), which we then correlated with the modularity index Q of the respective condition. One would expect a high magnitude of head motion to be inversely related with the modularity index. Across thresholds no significant associations between framewise displacement and modularity index could be observed (**Table S2**). This finding strongly suggests that modularity-differences across conditions are not the result of increased head motion during specific conditions.

**Table S2. Correlation between framewise displacement and modularity index for each of the six conditions across all six density thresholds**

| Condition (Q, FD) | 5% | | 10% | | 15% | | 20% | | 25% | | 30% | |
|---|---|---|---|---|---|---|---|---|---|---|---|---|
| | ***r*** | ***p*** | ***r*** | ***p*** | ***r*** | ***p*** | ***r*** | ***p*** | ***r*** | ***p*** | ***r*** | ***p*** |
| **Pre-task rest** | -.14 | .25 | -.06 | .66 | -.02 | .85 | .03 | .83 | .05 | .67 | .11 | .40 |
| **Reward** | -.25 | .09 | -.28 | .06 | -.22 | .14 | -.20 | .19 | -.08 | .58 | -.08 | .60 |
| **Reward+1** | -.27 | .06 | -.27 | .06 | -.26 | .08 | -.19 | .19 | -.15 | .31 | -.10 | .50 |
| **FNR** | -.25 | .10 | -.25 | .10 | -.27 | .07 | -.29 | .06 | -.21 | .18 | -.21 | .18 |
| **FNR+1** | -.10 | .47 | .04 | .74 | .11 | .41 | .15 | .25 | .19 | .17 | .20 | .13 |
| **Post-task rest** | -.06 | .62 | .18 | .16 | .24 | .05 | .21 | .09 | .19 | .14 | .19 | .13 |

**Abbreviations:** ***FD,*** framewise displacement; ***p,*** p-value; ***Q,*** modularity index; ***r,*** Pearson correlation coefficient
